# Prognostic value of admission H3.1 nucleosome levels in sepsis-associated acute kidney injury: a secondary analysis of the SISPCT randomised clinical trial

**DOI:** 10.1101/2025.01.28.25321238

**Authors:** Caroline Neumann, Frank Bloos, Teddy Tun Win Hla, Thomas Bygott, Holger Bogatsch, Michael Kiehntopf, Friedemann Börner, Adrian T Press, Michael Bauer, Andrew Retter, SepNet Critical Care Trials Group

## Abstract

**Background:** NETosis is a key innate immune defence mechanism where neutrophils release extracellular traps (NETs). However, excessive NET formation may damage organs during sepsis. We investigated the association between NETs and sepsis outcomes, including mortality and acute kidney injury (AKI).

**Methods:** We analysed levels of H3.1 nucleosomes in 971 patients with severe sepsis and septic shock from the SISPCT trial (Effect of Sodium Selenite Administration and Procalcitonin-Guided Therapy on Mortality in Patients With Severe Sepsis or Septic Shock). We evaluated associations between H3.1 levels and mortality and the need for renal replacement therapy using multivariable Cox regression and receiver operating characteristic analyses.

**Results:** We analysed 971 critically ill patients with complete data including admission H3.1 levels. 443 patients (45.6%) presented with sepsis, 520 (53.6%) had septic shock, and eight patients had an unknown diagnosis as defined by Sepsis-3. Admission H3.1 levels were higher in patients with septic shock than with sepsis (median 921.84 vs 432.71 ng/mL; p<0.001). Admission H3.1 levels were higher in non-survivors, and in a univariate Cox analysis, each log-10 increase in H3.1 was associated with a hazard ratio of 1.86 (95% confidence interval 1.41–2.47, p<0.05). H3.1 was also higher in patients requiring renal replacement therapy with septic shock vs sepsis (1832 ng/mL vs 801.4 ng/mL, p=0.01) and demonstrated a dose-response relationship with the severity of AKI.

**Conclusion:** Elevated levels of H3.1 nucleosomes at admission are independently associated with mortality and severe kidney dysfunction requiring renal replacement therapy.

**Trial registration:** Clinicaltrials.gov Identifier, NCT00832039.

## Introduction

Sepsis remains a massive global health challenge, affecting approximately 48.9 million people annually and causing 11 million deaths worldwide [1]. Sepsis is characterised by a dysregulated host response to infection [2]. It frequently leads to acute kidney injury (AKI), with an estimated 19–33% of intensive care patients developing sepsis-associated AKI (SA-AKI) [3,4]. Whilst no universal definition of SA-AKI exists, we follow the international consensus definition of the presence of both Sepsis-3 criteria and AKI using Kidney Disease Improving Global Outcomes (KDIGO) criteria [5]. This complication significantly increases mortality from 30% to 45% [6], with rates exceeding 60% when renal replacement therapy (RRT) is required [7]. Long-term outcomes are equally concerning, with 20–30% of SA-AKI survivors progressing to chronic kidney disease (CKD) within 3–5 years [8].

Early identification of SA-AKI could allow for more targeted interventions and improve outcomes. These challenges underscore the urgent need for novel biomarkers to identify and characterise subtypes of sepsis, sepsis trajectories and sepsis-associated organ dysfunctions more effectively.

Neutrophil extracellular traps (NETs) were first reported in 2004 [9]. H3.1 nucleosome release is reported as a general marker of NETosis and it has emerged as a potential biomarker candidate [10,11]. There has been growing interest in the role of extracellular histones and nucleosomes in the pathophysiology of sepsis [12,13]. During the innate immune response, histones are released extracellularly, either freely or within nucleosomes, functioning as damage-associated molecular patterns. These molecules exhibit cytotoxic and pro-inflammatory properties, contributing to endothelial dysfunction, coagulation abnormalities and organ injury [14]. Elevated circulating histone levels correlate with disease severity in meningococcal sepsis [15]. Experimental studies demonstrate that histone injection induces AKI in animal models [16]. The pathogenic mechanisms include direct renal tubular toxicity, pro-inflammatory cytokine induction and promotion of intravascular coagulation [17].

Recent research indicates that nucleosomes may serve as more stable and specific sepsis biomarkers than free histones, with documented correlations between nucleosome levels and disease severity and mortality [18]. The role of H3.1 nucleosomes in patients presenting with sepsis and SA-AKI has not yet been studied in a large sample of critically unwell patients. This study utilised samples from the SISPCT (Effect of Sodium Selenite Administration and Procalcitonin-Guided Therapy on Mortality in Patients With Severe Sepsis or Septic Shock) randomised clinical trial to evaluate the potential of H3.1 nucleosome clinical utility as a biomarker for prediction of sepsis outcomes and severe kidney dysfunction.

## Methods

### Aims

This analysis has four aims:

1. To quantify the admission H3.1 nucleosome levels in patients with sepsis and septic shock
2. To investigate the correlation between admission H3.1 nucleosome levels with admission white cell count (WCC) and organ severity scores
3. To determine the predictive values of plasma H3.1 nucleosome levels between admission and 14-day, 28-day and 90-day mortality
4. To investigate the admission plasma H3.1 nucleosome levels as a biomarker for RRT.

### Study design and study population

We conducted a secondary analysis of circulating H3.1 nucleosomes in plasma samples collected from patients with sepsis and septic shock who participated in the multicentre, randomised, bifactorial, prospective SISPCT trial (Clinicaltrials.gov identifier, NCT00832039) [19]. The study design and population has been described previously in detail and full eligibility criteria are provided in the *Supplemental Methods* [19]. The trial was conducted from 6 November 2009 to 6 June 2013, and included a 90-day follow-up period.

This secondary analysis from the SISPCT trial was accepted by the ethical committee of Friedrich-Schiller University Jena, Germany on 4 April 2023 (registration number: 2023– 2937–Material).

### Blood sample and data collection

Blood samples were collected on days 0 to 10, and day 14 after randomisation if the patient was still in the intensive care unit (ICU). Citrate plasma samples were bio-banked in the Integrated Biobank Jena and stored at −80°C. All relevant data on demographics, comorbidities such as the Charlson Comorbidity Index, laboratory findings, diagnostic tests, and organ support in critically ill ICU patients (mechanical ventilation, administration of vasopressors and RRT), and time of admission or transfer were extracted, reviewed and recorded from the hospital database. Overall survival outcome was assessed for up to 90 days. In case of an earlier hospital discharge, patients received a follow-up call from study personnel as agreed on enrolment.

### Nucleosome measurements

According to the manufacturer’s instructions, H3.1 nucleosome concentrations were measured in frozen citrate plasma samples using Nu.Q^®^ NETs immunoassay (CE-IVDD, Belgian Volition SRL, Isnes, Belgium). The instructions are summarised in *Supplemental Appendix A*.

### Sepsis outcomes and kidney failure

We reported 7-day, 14-day, 28-day and 90-day mortality and used the need for RRT as a time-dependent outcome. AKI stages were defined according to the KDIGO guidelines [20] and we excluded patients who were already established on RRT in the analysis regarding AKI. We did not discriminate between continuous and intermittent modes of RRT, and participating hospitals delivered RRT using either continuous veno-venous haemodialysis or intermittent RRT. We considered RRT an objective endpoint for severe kidney failure. RRT was initiated based on absolute indications: severe electrolyte disturbances, persistent metabolic acidosis, fluid (diuretics resistant) overload or severe uraemia and subsequent complications.

### Statistical analysis

Distributions of each variable were reviewed as per the SISPCT trial statistical analysis plan [19]. Admission H3.1 nucleosome levels were compared in sepsis and septic shock groups and across baseline characteristics. Continuous numerical variables were summarised using median values and categorical variables as counts and percentages of total. The Mann– Whitney test was used to determine whether there was a difference between the distribution of values for patients who had septic shock versus sepsis.

Scatter plots and correlation analysis were undertaken to explore the relationship between admission H3.1 nucleosome levels and other continuous numeric variables. Univariate Cox proportional hazards regression model were employed against multiple time-to-mortality and time-to-first RRT outcomes, and hazard ratios (HR) and 95% confidence intervals (CIs) were reported. For survival analyses, patients were right censored at hospital discharge or last known follow-up if they were lost to follow-up before the analysed time point (7, 14, 28 or 90 days). Subsequently, receiver operating characteristic (ROC) curve analysis was undertaken with area under the curve (AUROC) and its 95% CIs were calculated. Optimal cut-off points were determined by leveraging maximally selected rank statistics using *surv_cutpoint* function from *survminer* R package and *maxstat* R package [https://cran.r-project.org/web/packages/survminer/index.html] [https://cran.r-project.org/web/packages/maxstat/index.html]. Please see *Supplemental Appendix E* for detailed methodology. All statistical analyses were performed using R 4.1.0 (R Core Team, Vienna, Austria). We followed the CONSORT guidelines for reporting [21].

## Results

### Study population

Plasma samples were available for analysis from 971 patients from the SISPCT trial. The baseline characteristics and clinical features are summarised in Table 1. In our cohort of 971 critically ill patients, 443 patients (45.6%) presented with sepsis, 520 patients (53.6%) had septic shock, and eight patients had an unknown diagnosis (due to missing lactate measurements) as defined by the Sepsis-3 definition [2]. The median age of patients was 68 years and the majority (65%) were male. The sex distribution, median body mass index, Charlson-Deyo comorbidity index and Sequential Organ Failure Assessment (SOFA) scores were comparable between the groups.

**Table 1.** Baseline demographics and clinical characteristics.

|  | Total n=971; eligible n=963 | Sepsis-3 definition |  | P value |
| --- | --- | --- | --- | --- |
|  | Median [1stQ; 3rdQ] | Sepsis n=443 | Septic shock n=520 | <sup>b</sup>Mann–Whitney |
| **Demographics and comorbidity score** |  |  |  |  |
| Age, years | 68 [57; 75] | 67 [56; 74] | 67 [58; 76] | 0.1264 |
| Male, n (%) | 631 (65%) | 295 (67%) | 330 (63%) | 0.3108 |
| Body mass index (kg/m<sup>2</sup>) | 26.47 [23.64; 30.71] | 27.02 [24.03; 30.93] | 26.12 [23.43; 30.11] | 0.0957 |
| Charlson-Deyo comorbidity index | 2.0 [1.0; 4.0] | 2.0 [1.0; 4.0] | 2.0 [1.0; 4.0] | 0.2187 |
| Total SOFA score | 10 [8; 12] | 9 [7; 11] | 10 [8; 13] | <0.001 |
| SOFA subscores |  |  |  |  |
| Circulatory | 4 [3; 4] | 4 [3; 4] | 4 [4; 4] | <0.001 |
| Pulmonary | 3 [2; 3] | 3 [2; 3] | 3 [2; 3] | 0.002 |
| Coagulation | 0 [0; 1] | 0 [0; 0] | 0 [0; 1] | <0.001 |
| Liver | 0 [0; 1] | 0 [0; 1] | 0 [0; 1] | <0.001 |
| Neurological | 0 [0; 2] | 0 [0; 2] | 0 [0; 2] | 0.6431 |
| Renal | 1 [0; 3] | 1 [0; 2] | 1 [0; 3] | <0.001 |
| SAPS II | 61 [54; 70] | 59 [50; 68] | 63 [56; 72] | <0.001 |
| APACHE II | 23 [19; 28] | 22 [17; 26.5] | 25 [20; 30] | <0.001 |
| PaO<sub>2</sub>/FiO<sub>2</sub> (mmHg) | 160 [110; 235] | 170 [120; 246.8] | 153 [104; 218] | 0.0008 |
| **Admission laboratory parameters** |  |  |  |  |
| Haemoglobin mmol/L | 5.84 [5.15; 6.89] | 5.84 [5.15; 6.90] | 5.82 [5.15; 6.81] | 0.7583 |
| Platelets 10<sup>9</sup>/L | 193 [124; 288] | 224 [155; 324] | 169 [107; 245] | <0.001 |
| White cell count, 10<sup>6</sup>/L | 15.15 [10.5; 21.2] | 15.2 [10.6; 20.85] | 15.3 [10.2; 21.8] | 0.9784 |
| Urea, mmol/L | 9.296 [5.81; 14.91] | 8.134 [5.00; 13.45] | 10.126 [6.47; 15.60] | <0.001 |
| Creatinine, µmol/L | 114.92 [79.6; 185.6] | 97.24 [70.7; 159.1] | 140.72 [88.6; 194.9] | <0.001 |
| Albumin, g/L | 19 [15; 23] | 20 [16; 24] | 19 [15; 23] | 0.0109 |
| Bilirubin, mg/dL | 11.97 [7.9; 22.2] | 10.26 [6.8; 18.8] | 14.4 [8.6; 27.4] | <0.001 |
| CRP, mg/L | 227 [155; 305] | 223.5 [147; 304] | 229 [161; 307] | 0.2954 |
| Procalcitonin, µg/L | 6.67 [1.57; 26.0] | 2.60 [0.79; 9.3] | 13.17 [4.6; 42.4] | <0.001 |
| **Biomarker of interest** |  |  |  |  |
| **Admission H3.1 levels, ng/mL** | 629.51 [257; 1595] | 432.71 [184; 963] | 921.84 [369; 2394] | <0.001 |
| **ICU admission sources [3]** |  |  |  |  |
| Emergency medicine | 412 (42.4%) | 179 (40.4%) | 231 (44.4%) |  |
| Emergency surgery | 449 (46.2%) | 201 (45.4%) | 242 (46.5%) |  |
| Elective surgery | 110 (11.3%) | 63 (14.2%) | 47 (9.0%) |  |
| Cardiac arrest | 33 (3.4%) | 6 (1.4%) | 26 (5.0%) |  |
| **Sources of infection** |  |  |  |  |
| Community acquired | 427 (44.0%) | 172 (38.8%) | 251 (48.3%) |  |
| Nosocomial ward | 312 (32.1%) | 138 (31.2%) | 171 (32.9%) |  |
| Nosocomial ITU | 232 (23.9%) | 133 (30.0%) | 98 (18.8%) |  |

Continuous variables including duration are presented as median and interquartile ranges, categorical variables are presented as absolute value and percentage unless otherwise stated; <sup>b</sup>Mann–Whitney p value for sepsis versus septic shock population.
APACHE II, Acute Physiology and Chronic Health Evaluation II; CRP, C-reactive protein; ICU, intensive care unit; ITU, intensive treatment unit; Q, quartile; SAPS II, Simplified Acute Physiology Score II; SOFA, Sequential Organ Failure Assessment.

Admission H3.1 nucleosome levels were higher in patients with septic shock compared to patients with sepsis (median values 921.84 vs 432.71 ng/mL, p<0.001) (Table 1). Table 2 summarises the comparison of admission H3.1 nucleosome levels between the sepsis and septic shock populations according to a patient’s comorbidities. Table 3 summarises H3.1 nucleosome levels across complications of sepsis, interventions and mortality. Overall, the highest levels of nucleosomes were detected in the septic shock population across various diagnoses, sources of infection and hospital admission statuses. Pneumonia, at 29.4%, was the most common respiratory diagnosis, with significantly higher H3.1 nucleosome levels among those affected in the septic shock group versus sepsis group (1056 vs 407.7 ng/mL, p<0.001). With regards to AKI, a dose-response relationship was observed with the highest admission H3.1 levels in AKI 3 and RRT groups compared to patients without AKI.

**Table 2.** Admission H3.1 levels across comorbidities.

|  | Total, n (%) | Median [1st quartile; 3rd quartile]<br>admission H3.1 values, ng/mL |  |  | Sepsis-3 definition<br>Number of patients |  |  |
| --- | --- | --- | --- | --- | --- | --- | --- |
|  |  | Sepsis | Septic shock | P value | Sepsis | S shock | Missing |
|  | 971 | 432.7 | 921.8 | <.0001 | 443 | 520 | 8 |
| **Comorbidities** |  |  |  |  |  |  |  |
| Respiratory | 392 (40.4%) | 405.3 [177.3; 956.1] | 1041 [353.3; 2,725] | <0.001 | 182 (41.1%) | 209 (40.2%) | 1 |
| Neoplasms | 290 (29.9%) | 475.5 [196.7; 841.6] | 888.3 [323.9; 2,062] | 0.0004 | 127 (28.7%) | 162 (31.2%) | 1 |
| Circulatory | 282 (29.0%) | 462.6 [177.4; 1063] | 1066 [397.4; 2038] | <0.001 | 136 (30.7%) | 143 (27.5%) | 3 |
| Neurological | 123 (12.7%) | 334.7 [176.7; 740.6] | 971.4 [367.3; 2667] | 0.0009 | 58 (13.1%) | 65 (12.5%) | 0 |
| Rheumatological | 9 (0.9%) | 201.1 [177.9; 224.3] | 1120 [365.3; 1784] | NA | 3 (0.7%) | 6 (1.2%) | 0 |
| Gastrointestinal | 188 (19.4%) | 637 [171.1; 1,335] | 1081 [401; 2613] | 0.0068 | 67 (15.1%) | 121 (23.3%) | 0 |
| Genitourinary | 90 (9.3%) | 460.2 [312.8; 852] | 1143 [485.4; 2457] | 0.0097 | 32 (7.2%) | 57 (11.0%) | 1 |
| Gynaecological or<br>obstetric | 13 (1.3%) | 330.7 [184.8; 389.3] | 2489 [448.3; 3241] | 0.0734 | 6 (1.4%) | 7 (1.3%) | 0 |
| Endocrine and<br>metabolic | 247 (25.4%) | 429.3 [184.2; 1,041] | 1102 [415.2; 3,532] | <0.001 | 108 (24.4%) | 139 (26.7%) | 0 |
| Immunocompromised | 27 (2.8%) | 391 [191.4; 744.1] | 1115 [630.4; 3840] | 0.008 | 12 (2.7%) | 15 (2.9%) | 0 |

NA, not available.

**Table 3.**
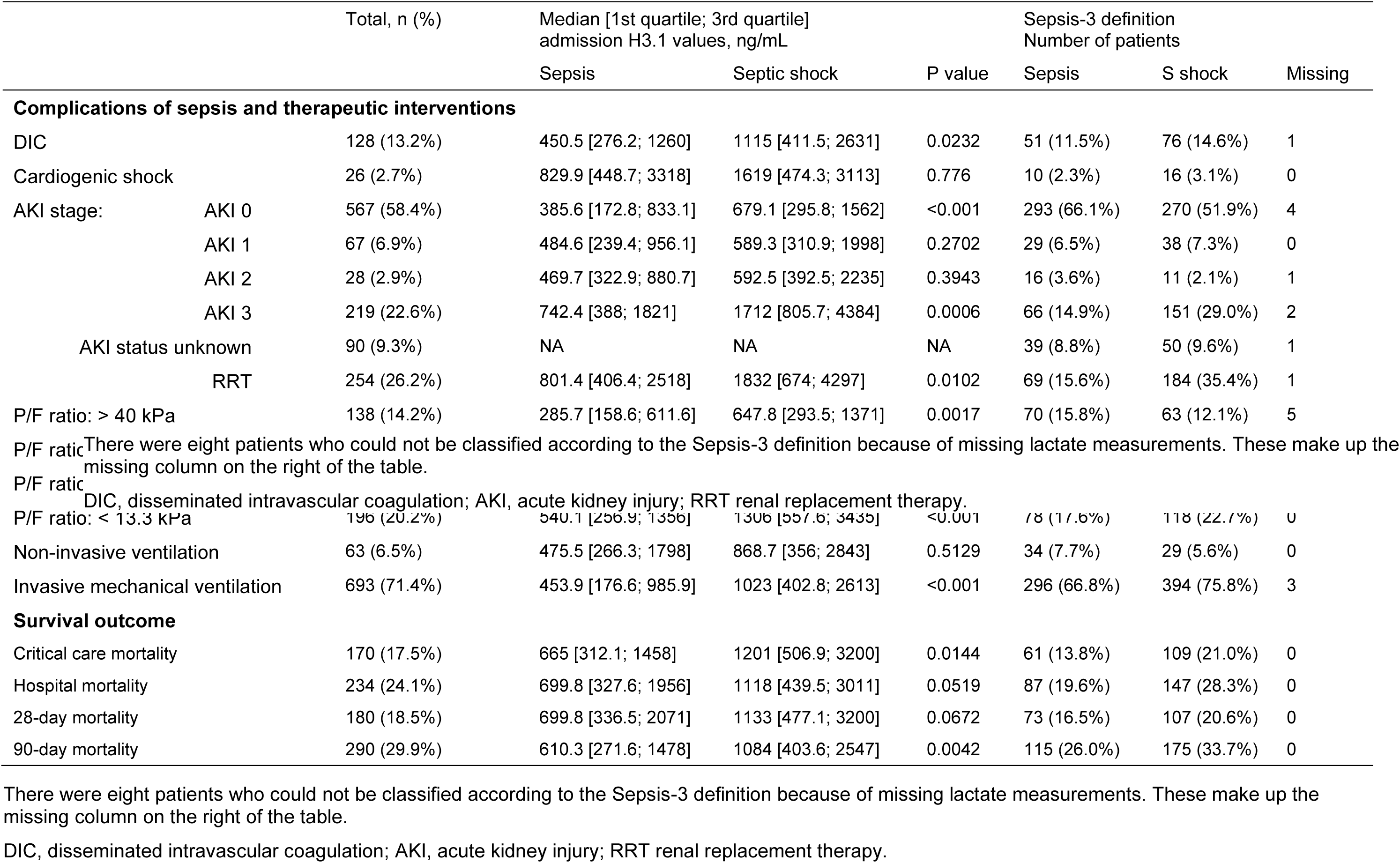
Admission H3.1 levels across therapeutic interventions and mortality outcomes.

### Correlation of H3.1 levels with WCC and organ severity scores

Admission H3.1 nucleosome levels showed a weak, statistically significant positive correlation with WCCs (Pearson correlation coefficient r=0.13 [0.06; 0.19], p<0.05), indicating likely independence between NET levels and WCCs. Further correlation analysis demonstrated a weak positive association between admission H3.1 nucleosomes and organ severity scores such as Acute Physiology and Chronic Health Evaluation II, SOFA and Simplified Acute Physiology Score II, and similarly with procalcitonin, C-reactive protein (CRP) and admission lactate (Supplemental Fig. 1 and Supplemental Table 1).

### Mortality

Overall, 73 (16.5%) out of 443 patients died in the sepsis group and 107 (20.6%) out of 520 patients died in the septic shock group within 28 days. For 90-day mortality, 115 (26%) out of 443 deaths were observed in the sepsis group and 175 (33.7%) out of 520 in the septic shock group.

In a univariate Cox model, log-10 transformed admission H3.1 levels were significantly associated with both 28-day and 90-day mortality outcomes (HR 1.86, 95% CI 1.41–2.47, p<0.05 and HR 1.54, 95% CI 1.24–1.93, p<0.05 respectively, Fig. 1). The 28-day mortality analysis revealed maximum lactate as the strongest predictor (HR 2.64, 95% CI 1.65–4.21, p=4.73e-05), closely followed by H3.1 (HR 1.86, 95% CI 1.41–2.47, p=1.37e-05).

**Fig. 1.**
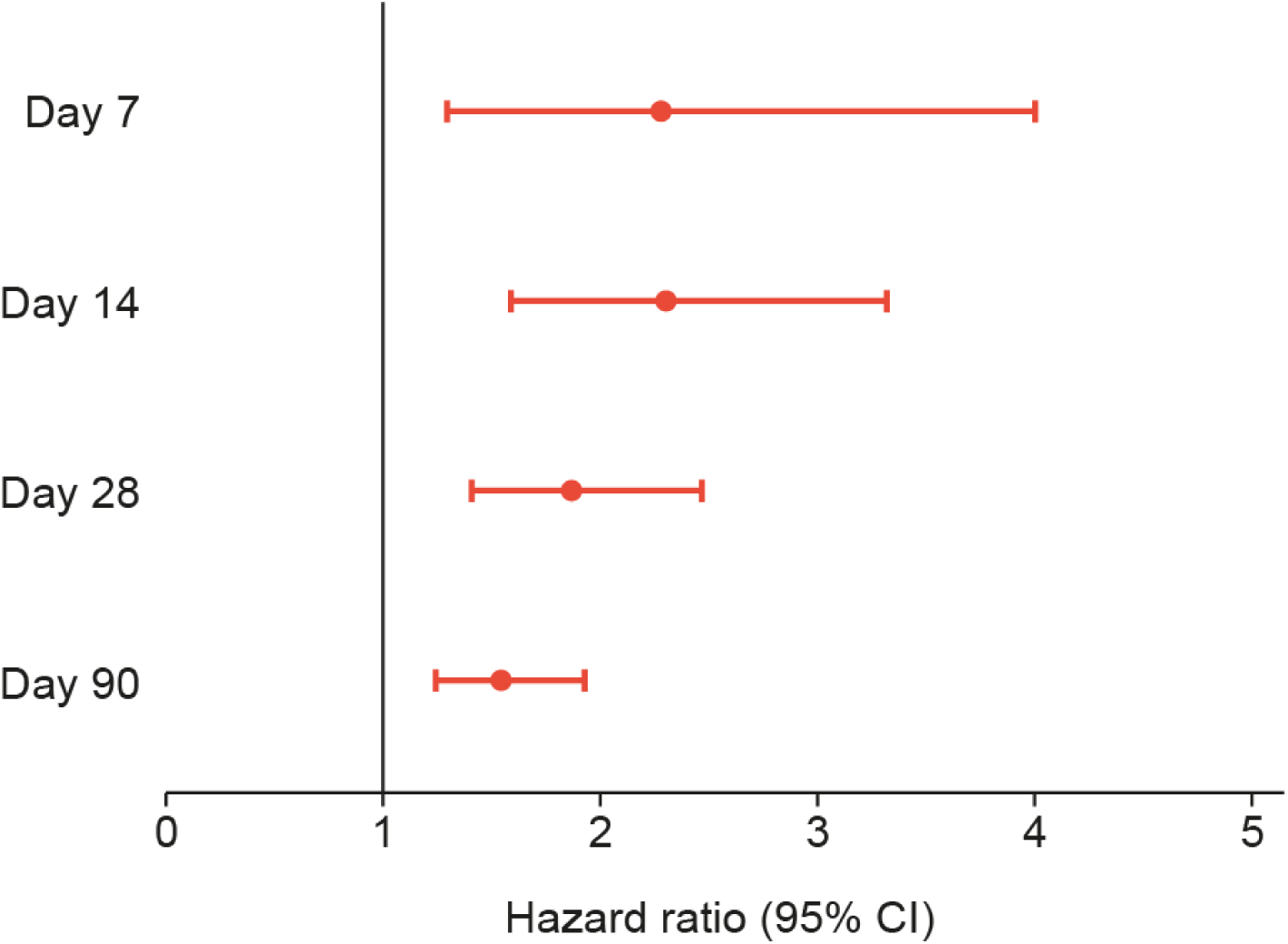
Association between log-transformed H3.1 nucleosome levels at admission and mortality risk over time in sepsis patients. Forest plot demonstrating the hazard ratios (HRs) with 95% confidence intervals (CIs) for mortality at days 7, 14, 28 and 90 in relation to admission H3.1 nucleosome levels (log-10 transformed) in sepsis and septic shock patients. The analysis shows the strongest association between H3.1 levels and early mortality (day 7: HR 2.28, 95% CI 1.30–4.01; day 14: HR 2.30, 95% CI 1.59–3.32), with a gradual attenuation over time (day 28: HR 1.86, 95% CI 1.41–2.47; day 90: HR 1.54, 95% CI 1.24– 1.93). All associations remained statistically significant (p<0.05) throughout the 90-day follow-up period. The raw data are shown in Supplemental Table 3.

**Fig. 2.**
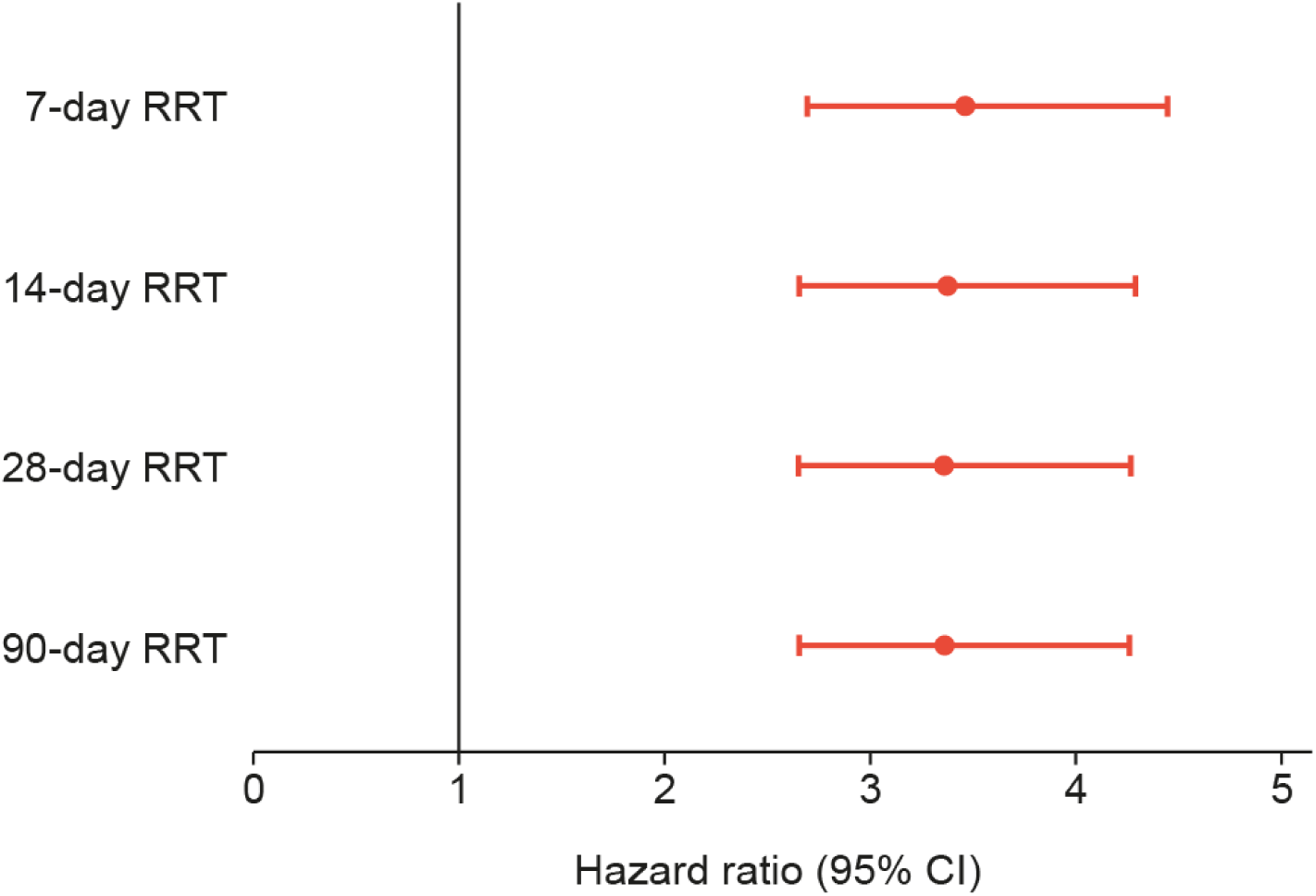
Forest plots of hazard ratios for RRT outcomes. Forest plot showing adjusted hazard ratios with 95% confidence intervals (CIs) for renal replacement therapy (RRT) outcomes at different time points after admission in sepsis and septic shock patients (n=831). The analysis demonstrates consistent hazard ratios of approximately 3.5–4.0 across all time periods (7, 14, 28 and 90 days), indicating that elevated admission H3.1 nucleosome levels (>2500 ng/mL) are associated with increased risk of requiring RRT. The sustained elevation in hazard ratios suggests that admission H3.1 levels may have prognostic value for predicting RRT requirements throughout the critical care stay. The raw data are shown in Supplemental Table 7.

Similarly, when we explored the admission H3.1 nucleosome levels across various time points for mortality, we found the strongest statistically significant association with mortality at day 7 (HR 2.28, 95% CI 1.30–4.01, p<0.05). Whilst this association attenuates over time, a statistically significant strong association persists at day 90 (HR 1.54, 95% CI 1.24–1.93, p<0.05)

Analysis of the ROC curves for five biomarkers: admission H3.1 levels, WCC, maximum lactate, CRP and procalcitonin, revealed varying predictive capabilities for survival outcomes across different time points (Supplemental Figs 2 to 6). The biomarkers demonstrate modest discriminatory power, with areas under the curve (AUC) ranging from 45% to 62%. H3.1 demonstrated peak performance at 14-day mortality with an AUC of 62.21% (95% CI 55.39– 69.03), whilst maintaining relatively stable predictive value across other time points. Maximum lactate showed comparable performance, with its highest AUC of 61.02% (95% CI 51.18–70.86) observed at day 7, followed by sustained performance through subsequent time points.

### Acute kidney injury

Of the 971 patients with plasma samples available, 33 patients were excluded due to pre-existing CKD in receipt of regular intermittent haemodialysis and 11 patients had missing RRT time series data, leaving 927 patients eligible for the 28-day RRT analysis. In this cohort of 927 patients, 216 required continuous RRT, six received intermittent dialysis and 705 did not require RRT. After removing all cases with missing values for H3.1 nucleosomes, platelets and urine output, a complete case analysis is available for 831 patients who experienced 198 continuous RRT, six intermittent dialysis and 627 no RRT (Supplemental Fig. 9 CONSORT diagram).

In patients who required RRT, median H3.1 nucleosome levels were higher in patients with septic shock (Table 2). In a univariate Cox model, log-10 transformed admission H3.1 levels were significantly associated with time-to-first RRT (HR 3.36, 95% CI 2.65–4.26, p<0.05).

On ROC analysis for RRT, admission H3.1 levels demonstrated moderate discriminatory performance with an AUC of 70.38% (95% CI 66.13–74.64). When compared with other individual biomarkers, H3.1’s performance was comparable but not superior to traditional markers such as serum creatinine and urine output. Initial creatinine level showed the strongest individual predictive capability with an AUC of 78.31% (95% CI 75.01–81.6), whilst initial lactate level (AUC = 67.65%, 95% CI 63.56–71.74) and CRP (AUC = 54.39%, 95% CI 49.79–58.98) demonstrated lower predictive value (Supplemental Fig. 11). After determining an optimal cut-off using rank statistics (as described above), we identified a cut-off value of admission H3.1 of 2500 ng/mL and above this threshold, the HR for a patient requiring RRT was 4.07 (CI 3.06–5.4). Finally, a clinical model was produced using a combination of H3.1 nucleosomes, platelets and 24-hour urine output (Supplemental Fig. 12). Patients with the highest score (Score 3) showed markedly worse outcomes, with only about 5% avoiding RRT by day 28, while those with Score 0 had over 90% RRT-free survival. Clear stratification was observed across all risk groups, suggesting strong discriminative ability of this combined model.

## Discussion

In this secondary analysis of the SISPCT trial with 971 patients, we demonstrated that admission H3.1 nucleosome levels have prognostic value in patients with sepsis and septic shock, particularly for early mortality and need for RRT. This is the first observation from a large multicentre clinical trial evaluating the role of H3.1 in sepsis and SA-AKI.

NETs are essential to pathogen clearance. In excessive formation, NETs are able to encourage further destructive signalling through interaction with other tissue components and the immune system [22]. Antimicrobial histones, constituents within the NETs structure, impose a direct cytotoxic effect on tissues [23]. To date, there have been numerous accounts of NETosis as ‘inflammatory cell death mechanisms’ being present in diseases of major organs [22]. First, we observed significantly elevated H3.1 nucleosome levels in patients with septic shock compared to those with sepsis alone across all primary diagnosis categories (Table 1 and 2). This marked difference suggests that nuclear DNA released during innate immune response can contribute to remote tissue injury, exacerbates the extent of organ damage and correlates with disease severity.

Our finding aligns with studies in smaller cohorts of critically unwell patients where elevated circulating histones were observed in severe sepsis and septic shock, though this is the first study to specifically quantify H3.1 nucleosomes in a large multicentre cohort of sepsis patients across different diagnosis categories [24].

Novel insights into the involvement of NETosis in different organ injuries and potential therapeutic measures targeting NETosis remain largely unexplored areas. The prognostic value of admission H3.1 nucleosome levels was most pronounced in the early period following ICU admission, with HRs for mortality highest at 7 and 14 days (HR 2.28 and 2.30, respectively), with a gradual attenuation of effect over time. This temporal pattern suggests that nucleosome levels may be useful for identifying patients at the highest risk of early deterioration. Notably, patients with very high admission levels (>20,000 ng/mL) exhibited 100% mortality within 14 days, while those with levels below 1000 ng/mL had significantly better survival (93% survived). To our knowledge, these are some of the highest measurements of NETs observed, and this stratification suggested a potential role in identifying patients requiring more aggressive early intervention.

When compared with conventional biomarkers, H3.1 nucleosomes demonstrated comparable or superior prognostic performance for 28-day mortality. While maximum lactate showed a stronger association with 28-day mortality (HR 2.64 vs 1.86), H3.1 nucleosomes outperformed CRP and provided independent predictive value. However, our modelling using log-10 transformed H3.1 levels allows higher granularity than lactate levels. This granularity suggests that nucleosome measurements are from an independent biological pathway from lactate synthesis and could complement existing risk stratification tools, potentially adding value through its mechanistic link to cell death and inflammation.

Notably, in contrast to previous studies [24], H3.1 nucleosome levels showed only weak correlation with WCC (Pearson r=0.13), suggesting processes beyond ‘simple leukocytosis’. Similarly, this strengthens the use of NETs as a complementary biomarker. While serum creatinine levels at admission could represent AKI or CKD, the weak correlation with initial creatinine (Pearson r=0.09) but strong association with subsequent RRT suggests nucleosomes may indicate evolving organ dysfunction rather than just current injury. A simple comparison of H3.1 and serum creatinine in AKI prediction leads to a circular conclusion given serum creatinine as a predictor and as an AKI definition, with published literature addressing the shortcomings of using creatinine or urine output [25]. For this reason, we decided to evaluate the prediction of RRT as a clear maximal extent of the highest AKI severity using H3.1 levels. Patients requiring RRT had significantly higher nucleosome levels, particularly in the septic shock group (Table 2).

Moderate AUROC of 70.38% of H3.1 for RRT is comparable to urine and blood neutrophil gelatinase-associated lipocalin (NGAL) (AUC = 0.720 [95% CI 0.638–0.803] and 0.755 [95% CI 0.706–0.803], respectively) for prediction of RRT use in critically unwell patients [26]. However, the ideal circumstances for whether and when to start RRT remain unclear and recent studies demonstrated no survival benefit at day 90 in an accelerated renal-replacement strategy versus standard strategy [27]. However, we believe the strength and utility of H3.1 measurement is not in its role as an AKI biomarker but more in its role as a dysfunction of innate immune response causing organ injury. A study by He on 136 SA-AKI patients demonstrated that plasma NET markers may independently predict 28-day mortality in this cohort [28]; thus, 28-day mortality may be mediated by NET levels as a marker of organ dysfunction through RRT.

The study has several strengths: firstly, its large sample size, multicentre design, and comprehensive clinical characterisation and extensive exploration with mortality. The use of standardised H3.1 nucleosome measurement and predefined clinical endpoints enhances reliability. However, as a secondary analysis, the findings are hypothesis-generating and require prospective validation. Secondly, we only analysed admission nucleosome levels; serial measurements might provide additional prognostic information. Thirdly, while associations with outcomes are clear, the precise mechanisms linking nucleosome levels to organ dysfunction remain to be fully elucidated. Finally, in this study, no other AKI biomarkers were evaluated, such as damage biomarkers, e.g. kidney injury molecule-1 [29], NGAL or L-type fatty acid-binding protein [30], C–C motif chemokine ligand 14 [7,31] stress biomarkers, e.g. tissue inhibitor of metalloprotease-2 and insulin-like growth factor-binding protein-7 [32], and functional markers, e.g. proenkephalin A 119-159 [33], and we thus recommend future studies to evaluate such biomarkers of (sub-)clinical renal damage.

Our findings have clinical implications; first, the early prognostic value of H3.1 nucleosomes could help identify high-risk patients requiring more intensive monitoring or aggressive intervention. Second, the association with RRT might guide the timing of renal replacement initiation, though this requires prospective evaluation. Additionally, nucleosome levels could serve as a stratification tool for future clinical trials in sepsis and SA-AKI.

Future studies should evaluate serial nucleosome measurements to characterise their dynamic changes during sepsis progression. Investigating potential therapeutic implications, such as nucleosome-targeted interventions or the timing of RRT initiation based on nucleosome levels, may yield clinical benefits. Additionally, exploring the relationship between nucleosome levels and long-term outcomes, including CKD risk, would be valuable.

## Conclusion

Our study establishes H3.1 nucleosomes as a biomarker delineating organ dysfunction in sepsis, particularly for early mortality and RRT. The clear stratification of risk and independence from conventional markers suggests clinical utility, though prospective validation is needed. These findings advance our understanding of nucleosome biology in critical illness and help inform future therapeutic strategies.

## List of abbreviations

AKI: Acute kidney injury
APARHE II: Acute Physiology and Chronic Health Evaluation II
AUROC: Area under the receiver operating characteristic curve
AUC: Area under the curve
CI: Confidence interval
CKD: Chronic kidney disease
CRP: C-reactive protein
DIC: Disseminated intravascular coagulation
HR: Hazard ratio
ICU: Intensive care unit
ITU: Intensive treatment unit
KDIGO: Kidney Disease Improving Global Outcomes
NA: Not applicable
NET: Neutrophil extracellular traps
NGAL: Neutrophil gelatinase-associated lipocalin
Q: Quartile
ROC: Receiver operating characteristic
RRT: Renal replacement therapy
SA-AKI: Sepsis-associated acute kidney injury
SAPS II: Simplified Acute Physiology Score II
SOFA: Sequential Organ Failure Assessment
WCC: White cell count

## Acknowledgements

We would like to thank the patients who participated in this study. We thank all members of the SepNet Critical Care Trials Group: Thorsten Brenner, Essen; Patrick Meybohm, Würzburg; Josef Briegel, München; Markus Weigand, Heidelberg; Matthias Gründling, Greifswald; Holger Bogatsch, Leipzig; Markus Löffler, Leipzig; Gunnar Elke, Kiel; Sandra Frank, Munich; Melanie Meersch-Dini, Münster; Christian Putensen, Bonn; Achim Kaasch, Magdeburg, Stefan Kluge, Hamburg; all Germany. Medical writing support under author guidance, was provided by Georgina Collett, PhD, on behalf of Sparked into Life Ltd, Macclesfield, UK and funded by Volition Diagnostics UK Ltd, London, UK in accordance with Good Publication Practice (GPP 2022) Guidelines. Illustrations were created by Jim Park of Sparked into Life Ltd, Macclesfield, UK in collaboration with Dr Andrew Retter and funded by Volition Diagnostics UK Ltd, London, UK. The manuscript was edited by Michelle Thorpe on behalf of Sparked into Life Ltd, Macclesfield, UK and funded by Volition Diagnostics UK Ltd, London, UK. Ultimate responsibility for opinions, conclusions and data interpretation lies with the authors.

## Author contributions

Andrew Retter was responsible for conceptualising the study, analysing the initial data, and writing the first draft of the study. Teddy Tun Win Hla was responsible for secondary data analysis and contributed extensively to the write up of the manuscript. Thomas Bygott was the lead statistician responsible for supervising and enabling the data analysis. Michael Bauer assisted in the conceptualisation of the study and hypothesis generation. Caroline Neumann contributed to the data analysis and writing of multiple drafts of the manuscript. All author authors read and approved the final manuscript for publication.

## Funding

Deutsche Forschungsgemeinschaft (DFG) Project No. 542813223 and 316213987/B08, C06.

German Federal Ministry of Education and Research, Project No. 03RU2U071H.

The H3.1 nucleosome ELISAs were provided by Volition Diagnostics UK Ltd. Medical writing, editing and illustrations were funded by Volition Diagnostics UK Ltd.

## Data availability

Data availability is settled by the original trial mentioned in reference 19.

## Declarations

## Conflicts of interest

Andrew Retter is the Chief Medical Officer of Volition Diagnostics UK Ltd. He also holds shares in VolitionRx Limited. Thomas Bygott is an employee of Volition Diagnostics UK Ltd and holds shares in VolitionRx Limited. Teddy Tun Win Hla has received fees for consultancy to Volition Diagnostics UK Ltd. Michael Bauer is a member of the International Sepsis Forum and has served in addition on advisory boards for Volition, ThermoFisher/ B.R.A.H.M.S, ArtCline and Bayer Inc. All other authors declare no conflicts of interest.

Volition has patents covering Nu.Q^®^ technology and are developers of Nu.Q^®^ assays.

## Notes

### Author Declarations

This secondary analysis from the SISPCT trial was accepted by the ethical committee of Friedrich Schiller University Jena, Germany on 4 April 2023 (registration number: 2023 to 2937).

## References

1. Rudd KE, Johnson SC, Agesa KM, Shackelford KA, Tsoi D, Kievlan DR, et al. Global, regional, and national sepsis incidence and mortality, 1990–2017: analysis for the Global Burden of Disease Study. Lancet 2020;395:200–11. 10.1016/S0140-6736(19)32989-7

2. Singer M, Deutschman CS, Seymour CW, Shankar-Hari M, Annane D, Bauer M, et al. The Third International Consensus Definitions for Sepsis and Septic Shock (Sepsis-3). JAMA. 2016;315:801.

3. White KC, Serpa-Neto A, Hurford R, Clement P, Laupland KB, See E, et al. Sepsis-associated acute kidney injury in the intensive care unit: incidence, patient characteristics, timing, trajectory, treatment, and associated outcomes. A multicenter, observational study. Intensive Care Med. 2023;49:1079–89.

4. Peerapornratana S, Manrique-Caballero CL, Gómez H, Kellum JA. Acute kidney injury from sepsis: current concepts, epidemiology, pathophysiology, prevention and treatment. Kidney Int. 2019;96:1083–99.

5. Zarbock A, Nadim MK, Pickkers P, Gomez H, Bell S, Joannidis M, et al. Sepsis-associated acute kidney injury: consensus report of the 28th Acute Disease Quality Initiative workgroup. Nat Rev Nephrol. 2023;19:401–17.

6. Sakr Y, Lobo SM, Moreno RP, Gerlach H, Ranieri VM, Michalopoulos A, et al. Patterns and early evolution of organ failure in the intensive care unit and their relation to outcome. Crit Care. 2012;16:R222.

7. Hoste EAJ, Bagshaw SM, Bellomo R, Cely CM, Colman R, Cruz DN, et al. Epidemiology of acute kidney injury in critically ill patients: the multinational AKI-EPI study. Intensive Care Med. 2015;41:1411–23.

8. Forni LG, Darmon M, Ostermann M, Oudemans-van Straaten HM, Pettilä V, Prowle JR, et al. Renal recovery after acute kidney injury. Intensive Care Med. 2017;43:855–66.

9. Brinkmann V, Reichard U, Goosmann C, Fauler B, Uhlemann Y, Weiss DS, et al. Neutrophil extracellular traps kill bacteria. Science. 2004;303:1532–5.

10. Zukas K, Cayford J, Serneo F, Atteberry B, Retter A, Eccleston M, et al. Rapid high-throughput method for investigating physiological regulation of neutrophil extracellular trap formation. J Thromb Haemost 2024;22:2543–54. 10.1016/j.jtha.2024.05.028

11. Cayford J, Eccleston M, Berman B, Serneo F, Kelly T, Retter A, et al. Understanding NETosis priming and induction through nucleosome changes and real-time monitoring in isolated neutrophils and in an ex-vivo model. Blood 2023;142:2542. 10.1182/blood-2023-178100

12. Abrams ST, Morton B, Alhamdi Y, Alsabani M, Lane S, Welters ID, et al. A novel assay for neutrophil extracellular trap formation independently predicts disseminated intravascular coagulation and mortality in critically ill patients. Am J Respir Crit Care Med 2019;200:869–80. 10.1164/rccm.201811-2111OC

13. Alhamdi Y, Abrams ST, Cheng Z, Jing S, Su D, Liu Z, et al. Circulating histones are major mediators of cardiac injury in patients with sepsis. Crit Care Med 2015;43:2094–103. 10.1097/CCM.0000000000001162

14. Xu J, Zhang X, Pelayo R, Monestier M, Ammollo CT, Semeraro F, et al. Extracellular histones are major mediators of death in sepsis. Nat Med. 2009;15:1318–21.

15. Zeerleder S, Stephan F, Emonts M, de Kleijn ED, Esmon CT, Varadi K, et al. Circulating nucleosomes and severity of illness in children suffering from meningococcal sepsis treated with protein C. Crit Care Med 2012;40:3224–9. 10.1097/CCM.0b013e318265695f

16. Allam R, Scherbaum CR, Darisipudi MN, Mulay SR, Hägele H, Lichtnekert J, et al. Histones from dying renal cells aggravate kidney injury via TLR2 and TLR4. J Am Soc Nephrol 2012;23:1375–88. 10.1681/ASN.2011111077

17. Nakazawa D, Kumar SV, Marschner J, Desai J, Holderied A, Rath L, et al. Histones and neutrophil extracellular traps enhance tubular necrosis and remote organ injury in ischemic AKI. J Am Soc Nephrol 2017;28:1753–68. 10.1681/ASN.2016080925

18. Ekaney ML, Otto GP, Sossdorf M, Sponholz C, Boehringer M, Loesche W, et al. Impact of plasma histones in human sepsis and their contribution to cellular injury and inflammation. Crit Care 2014;18:543. 10.1186/s13054-014-0543-8

19. Bloos F, Trips E, Nierhaus A, Briegel J, Heyland DK, Jaschinski U, et al. Effect of sodium selenite administration and procalcitonin-guided therapy on mortality in patients with severe sepsis or septic shock: a randomized clinical trial. JAMA Intern Med 2016;176:1266. 10.1001/jamainternmed.2016.2514

20. Khwaja A. KDIGO clinical practice guidelines for acute kidney injury. Nephron Clin Pract 2012;120:c179–c184. 10.1159/000339789

21. Schulz KF, Altman DG, Moher D, CONSORT Group. CONSORT 2010 statement: updated guidelines for reporting parallel group randomised trials. BMJ. 2010;340:c332.

22. Cahilog Z, Zhao H, Wu L, Alam A, Eguchi S, Weng H, et al. The role of neutrophil NETosis in organ injury: novel inflammatory cell death mechanisms. Inflammation 2020;43:2021–32. 10.1007/s10753-020-01294-x

23. Saffarzadeh M, Juenemann C, Queisser MA, Lochnit G, Barreto G, Galuska SP, et al. Neutrophil extracellular traps directly induce epithelial and endothelial cell death: a predominant role of histones. PLoS ONE 2012;7:e32366. 10.1371/journal.pone.0032366

24. Haem Rahimi M, Bidar F, Lukaszewicz A-C, Garnier L, Payen-Gay L, Venet F, et al. Association of pronounced elevation of NET formation and nucleosome biomarkers with mortality in patients with septic shock. Ann Intensive Care. 2023;13:102.

25. Schetz M, Schortgen F. Ten shortcomings of the current definition of AKI. Intensive Care Med. 2017;43:911–3.

26. Klein SJ, Brandtner AK, Lehner GF, Ulmer H, Bagshaw SM, Wiedermann CJ, et al. Biomarkers for prediction of renal replacement therapy in acute kidney injury: a systematic review and meta-analysis. Intensive Care Med. 2018;44:323–36.

27. STARRT-AKI Investigators, Canadian Critical Care Trials Group, Australian and New Zealand Intensive Care Society Clinical Trials Group, et al. Timing of initiation of renal-replacement therapy in acute kidney injury. N Engl J Med 2020;383:240–51. 10.1056/NEJMoa2000741

28. He J, Zheng F, Qiu L, Wang Y, Zhang J, Ye H, et al. Plasma neutrophil extracellular traps in patients with sepsis-induced acute kidney injury serve as a new biomarker to predict 28-day survival outcomes of disease. Front Med. 2024;11:1496966.

29. Tanase DM, Gosav EM, Radu S, Costea CF, Ciocoiu M, Carauleanu A, et al. The predictive role of the biomarker kidney molecule-1 (KIM-1) in acute kidney injury (AKI) cisplatin-induced nephrotoxicity. Int J Mol Sci 2019;20:5238. 10.3390/ijms20205238

30. Yuan S-M. Acute kidney injury after cardiac surgery: risk factors and novel biomarkers. Braz J Cardiovasc Surg 2019;34:352–60. 10.21470/1678-9741-2018-0212

31. Bagshaw SM, Al-Khafaji A, Artigas A, Davison D, Haase M, Lissauer M, et al. External validation of urinary C-C motif chemokine ligand 14 (CCL14) for prediction of persistent acute kidney injury. Crit Care 2021;25:185. 10.1186/s13054-021-03618-1

32. Meersch M, Schmidt C, Van Aken H, Martens S, Rossaint J, Singbartl K, et al. Urinary TIMP-2 and IGFBP7 as early biomarkers of acute kidney injury and renal recovery following cardiac surgery. PloS One. 2014;9:e93460.

33. Caironi P, Latini R, Struck J, Hartmann O, Bergmann A, Bellato V, et al. Circulating proenkephalin, acute kidney injury, and its improvement in patients with severe sepsis or shock. Clin Chem 2018;64:1361–9. 10.1373/clinchem.2018.288068

