## Supplementary Material for "Prognostic value of admission H3.1 nucleosome levels in sepsis-associated acute kidney injury: a secondary analysis of the SISPCT randomised clinical trial"

This supplementary material package provides the technical details, methodological specifications, and additional analyses that support the main findings presented in the manuscript while keeping the main text focused and readable.

### Prognostic value of admission H3.1 nucleosome levels in sepsis-associated acute kidney injury: a secondary analysis of the SISPCT randomised clinical trial

Caroline Neumann[1] (ORCID: 0000-0002-9242-6505), Frank Bloos[1] (ORCID: 0000-0002-0767-7941), Teddy Tun Win Hla[2,3] (ORCID: 0000-0001-9769-1618), Thomas Bygott[4] (ORCID: 0009-0004-7017-1929), Holger Bogatsch[5], Michael Kiehntopf[6], Friedemann Börner[6], Adrian T Press[1,7,8] (ORCID: 0000-0002-6089-6764), Michael Bauer[1] (ORCID: 0000-0002-1521-3514) and Andrew Retter[2,9,*] (ORCID: 0000-0001-7373-6404); SepNet Critical Care Trials Group

**Author details**

[1]Department of Anaesthesiology and Intensive Care Medicine, Jena University Hospital, Jena, Germany. [2]Critical Care, Guy's and St Thomas, London, UK. [3]University College London Institute of Health Informatics London, London, UK. [4]Volition Diagnostics UK Ltd, London, UK. [5]Institute for Medical Informatics, Statistics and Epidemiology Clinical Trial Centre, Leipzig University, Leipzig, Germany. [6]Institute for Clinical Chemistry and Laboratory Diagnostics, Jena University Hospital, Jena, Germany. [7]Jena University Hospital, Center for Sepsis Control and Care, Friedrich-Schiller-University Jena, Jena, Germany. [8]Friedrich Schiller University Jena, Medical Faculty, Jena, Germany. [9]School of Immunology and Microbial Sciences, King's College, London.

#### Appendix A: Detailed methodology for H3.1 nucleosome measurements using Nu.Q® NETs immunoassay

H3.1 nucleosome concentrations were measured in frozen citrate plasma samples using Nu.Q® NETs immunoassay (CE-IVDD, Belgian Volition SRL, Isnes, Belgium) according to the manufacturer's instructions. This sandwich immunoassay is based on chemiluminescence technology and was performed using the IDS-i10 automated analyser system (Immunodiagnostic Systems Ltd, Boldon, UK). Briefly, 50 µL of thawed citrate plasma were incubated with acridinium ester-labelled anti-nucleosome detection antibody before magnetic particle beads coated with the monoclonal anti-histone variant H3.1 were added. After a wash step, trigger solutions were added and the light emitted by the acridinium ester was measured via the luminometer system. The nucleosome concentration of each sample is automatically calculated by the immunoassay analyser using a four-parameter logistic curve. All samples were analysed in duplicate. If the sample was above the limit of quantification and the coefficient of variation (%CV) of the determined concentration was above 20%, the analysis was repeated. Samples with concentrations >1200 ng/mL were automatically diluted to 1:5 by the instrument; those with estimates of concentration >6000 ng/mL were manually pre-diluted 10 times before retesting.

#### Appendix B: SISPCT trial criteria

##### Inclusion criteria

1. Presence of severe sepsis or septic shock
2. Onset of severe sepsis or septic shock not longer than 24 hours ago
3. Age at least 18 years
4. Written informed consent of the patient or the legal representative

##### Exclusion criteria

1. Pregnant or lactating women
2. Fertile female patients (<2 years after last menstruation) without appropriate contraception during study participation
3. Participation in a clinical study within the last 30 days
4. Current participation in another study or clinical trial
5. Previous participation in this clinical trial
6. Selenium intoxication
7. No commitment to full patient support (i.e. do not resuscitate order)
8. Patient's death is considered imminent due to coexisting disease
9. Relationship to the study team (i.e. colleague, relative, employee)
10. Infection where guidelines recommend a longer duration of antimicrobial therapy: (1) infections with *Listeria* spp, *Legionella pneumophila*, *Pneumocystis jiroveci* or *Mycobacterium tuberculosis*; (2) viral or parasitic infections (haemorrhagic fever, malaria); (3) bacterial endocarditis, brain abscess, bone or deep soft tissue infections, (4) chronic local infections (osteomyelitis)
11. Severely immunocompromised patients (i.e. HIV-infection with CD4 count <200 cells/mm$^3$, neutropenic patients (<500 neutrophils/mm$^3$), or patients with immunosuppressive therapy after solid organ transplantation)

**Appendix C: EQUATOR network checklist**

### CONSORT 2010 checklist of information to include when reporting a randomised trial*

| Section/topic | Item no. | Checklist item | Reported on page no. |
|---|---|---|---|
| **Title and abstract** | | | |
| | 1a | Identification as a randomised trial in the title | Pg 1 |
| | 1b | Structured summary of trial design, methods, results, and conclusions (for specific guidance see CONSORT for abstracts) | Pg 2 |
| **Introduction** | | | |
| Background and objectives | 2a | Scientific background and explanation of rationale | Pg 3 |
| | 2b | Specific objectives or hypotheses | Pg 4 |
| **Methods** | | | |
| Trial design | 3a | Description of trial design (such as parallel, factorial) including allocation ratio | Pg 4 |
| | 3b | Important changes to methods after trial commencement (such as eligibility criteria), with reasons | n/a |
| Participants | 4a | Eligibility criteria for participants | Supp material pg 4 |
| | 4b | Settings and locations where the data were collected | Supp material pg 4 |
| Interventions | 5 | The interventions for each group with sufficient details to allow replication, including how and when they were actually administered | n/a |
| Outcomes | 6a | Completely defined pre-specified primary and secondary outcome measures, including how and when they were assessed | Pg 5 |
| | 6b | Any changes to trial outcomes after the trial commenced, with reasons | n/a |
| Sample size | 7a | How sample size was determined | n/a |
| | 7b | When applicable, explanation of any interim analyses and stopping guidelines | n/a |
| Randomisation: | | | |
| Sequence generation | 8a | Method used to generate the random allocation sequence | n/a |
| | 8b | Type of randomisation; details of any restriction (such as blocking and block size) | n/a |
| Allocation concealment mechanism | 9 | Mechanism used to implement the random allocation sequence (such as sequentially numbered containers), describing any steps taken to conceal the sequence until interventions were assigned | n/a |
| Implementation | 10 | Who generated the random allocation sequence, who enrolled participants, and who assigned participants to interventions | n/a |
| Blinding | 11a | If done, who was blinded after assignment to interventions (for example, participants, care providers, those assessing outcomes) and how | n/a |

| | | | |
|---|---|---|---|
| | 11b | If relevant, description of the similarity of interventions | Table 3 page 21 |
| Statistical methods | 12a | Statistical methods used to compare groups for primary and secondary outcomes | Pg 6 |
| | 12b | Methods for additional analyses, such as subgroup analyses and adjusted analyses | Supp material pg 8 |
| **Results** | | | |
| Participant flow (a diagram is strongly recommended) | 13a | For each group, the numbers of participants who were randomly assigned, received intended treatment, and were analysed for the primary outcome | Supp material pg 7 |
| | 13b | For each group, losses and exclusions after randomisation, together with reasons | Supp material pg 7 and 23 |
| Recruitment | 14a | Dates defining the periods of recruitment and follow-up | Pg 4 |
| | 14b | Why the trial ended or was stopped | Pg 4 |
| Baseline data | 15 | A table showing baseline demographic and clinical characteristics for each group | Pg 18 |
| Numbers analysed | 16 | For each group, number of participants (denominator) included in each analysis and whether the analysis was by original assigned groups | Pg 18 |
| Outcomes and estimation | 17a | For each primary and secondary outcome, results for each group, and the estimated effect size and its precision (such as 95% confidence interval) | Pg 22 and 23 |
| | 17b | For binary outcomes, presentation of both absolute and relative effect sizes is recommended | n/a |
| Ancillary analyses | 18 | Results of any other analyses performed, including subgroup analyses and adjusted analyses, distinguishing pre-specified from exploratory | n/a |
| Harms | 19 | All important harms or unintended effects in each group (for specific guidance see CONSORT for harms) | n/a |
| **Discussion** | | | |
| Limitations | 20 | Trial limitations, addressing sources of potential bias, imprecision, and, if relevant, multiplicity of analyses | Pg 9 and 10 |
| Generalisability | 21 | Generalisability (external validity, applicability) of the trial findings | Pg 9 and 10 |
| Interpretation | 22 | Interpretation consistent with results, balancing benefits and harms, and considering other relevant evidence | Pg 9 and 10 |
| **Other information** | | | |
| Registration | 23 | Registration number and name of trial registry | Pg 4 |
| Protocol | 24 | Where the full trial protocol can be accessed, if available | n/a |
| Funding | 25 | Sources of funding and other support (such as supply of drugs), role of funders | Pg 13 and 14 |

*We strongly recommend reading this statement in conjunction with the CONSORT 2010 Explanation and Elaboration for important clarifications on all the items. If relevant, we also recommend reading CONSORT extensions for cluster randomised trials, non-inferiority and equivalence trials, non-pharmacological treatments, herbal interventions, and pragmatic trials. Additional extensions are forthcoming: for those and for up to date references relevant to this checklist, see www.consort-statement.org.

#### Appendix D: CONSORT diagram 1

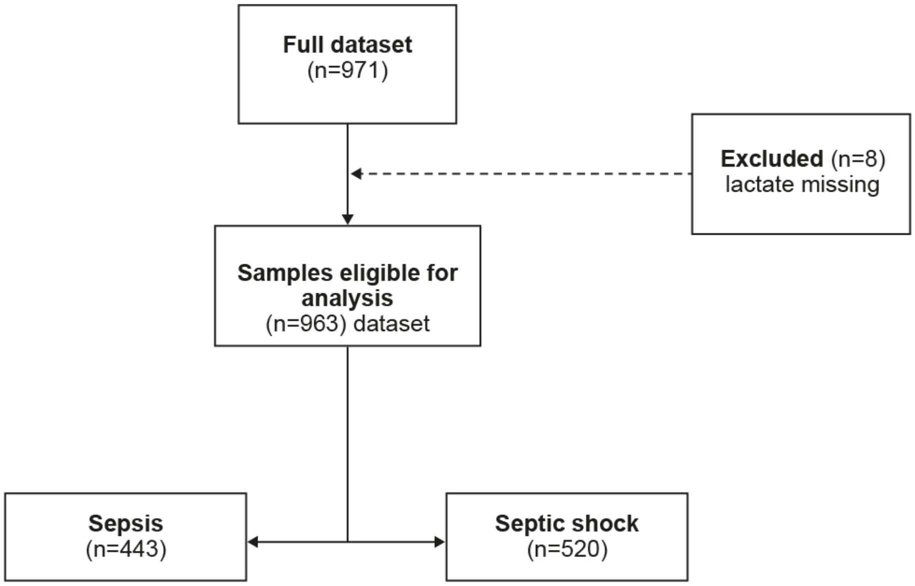

#### Appendix E: Methodology

##### Correlation analysis

Relationships between continuous variables were visualised using scatter plot and coefficient determined using Spearman coefficient.

##### ROC curve analysis

To evaluate the diagnostic performance of admission H3.1 nucleosome concentrations, the area under the receiver operating characteristic curve (AUROC) was calculated along with their 95% confidence intervals. The AUROC curves for admission H3.1 nucleosome levels were compared to admission white cell count, C-reactive protein (CRP), procalcitonin and the patient's maximum lactate concentration.

Thereafter, the optimal cut-off point to differentiate mortality for each of the variables was calculated using *surv_cutpoint* from the *survminer* R package, which uses maximally selected rank statistics. To create an ordering within these groups, patients were ranked by each variable in turn, starting with the most statistically significant variable when each cut-off point was calculated. For individual biomarkers this was done by creating a Boolean variable with 1 for values equal to or above the cut-off and 0 for those below and running coxph afterwards.

##### Cox model

Cox proportional hazards regression was employed and admission H3.1 nucleosome levels were compared to admission white cell count, CRP, procalcitonin and the patient's maximum lactate concentration. Proportional hazards assumptions were checked and visually verified.

#### Appendix F: Correlation between H3.1 and other variables

**Fig. 1** Correlation between admission H3.1 nucleosome levels and white cell count

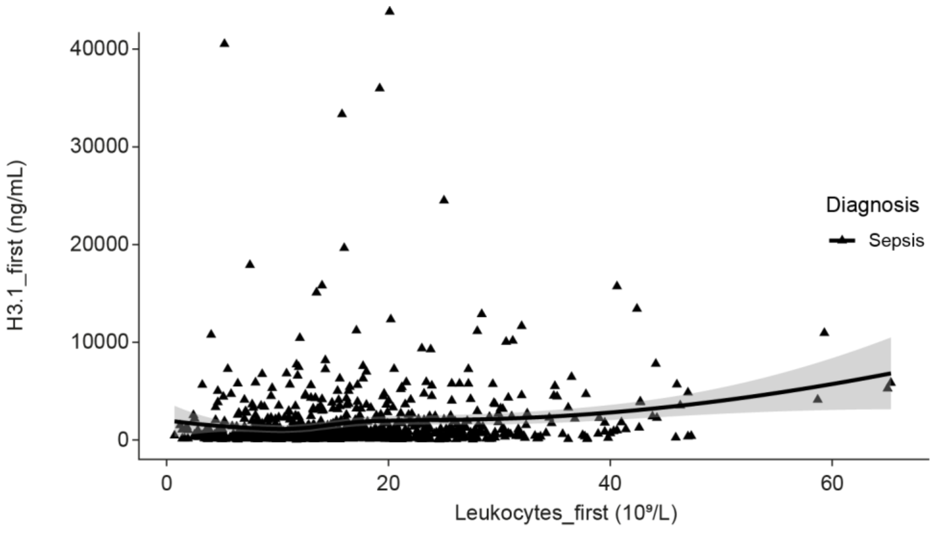

**Table 1** Spearman correlation coefficients between H3.1 and baseline variables

| Baseline variable | Spearman correlation, % |
|---|---|
| **Demographics** | |
| Sex | 11 |
| Age | 5 |
| Weight | 4 |
| Height | -11 |
| Body mass index | 10 |
| **Scoring system** | |
| APACHE II | 26 |
| SOFA | 27 |
| SAPS II | 22 |
| **Biomarkers** | |
| CRP | 17 |
| Procalcitonin | 30 |
| Lactate | 30 |
| Leucocytes | 17 |

APACHE II, Acute Physiology and Chronic Health Evaluation II; CRP, C-reactive protein; SAPS II, Simplified Acute Physiology Score II; SOFA, Sequential Organ Failure Assessment.

#### Appendix G: Additional supplementary figures (2–6)

**Fig. 2** Initial H3.1 day 7, 14, 28 and 90 ROC curves

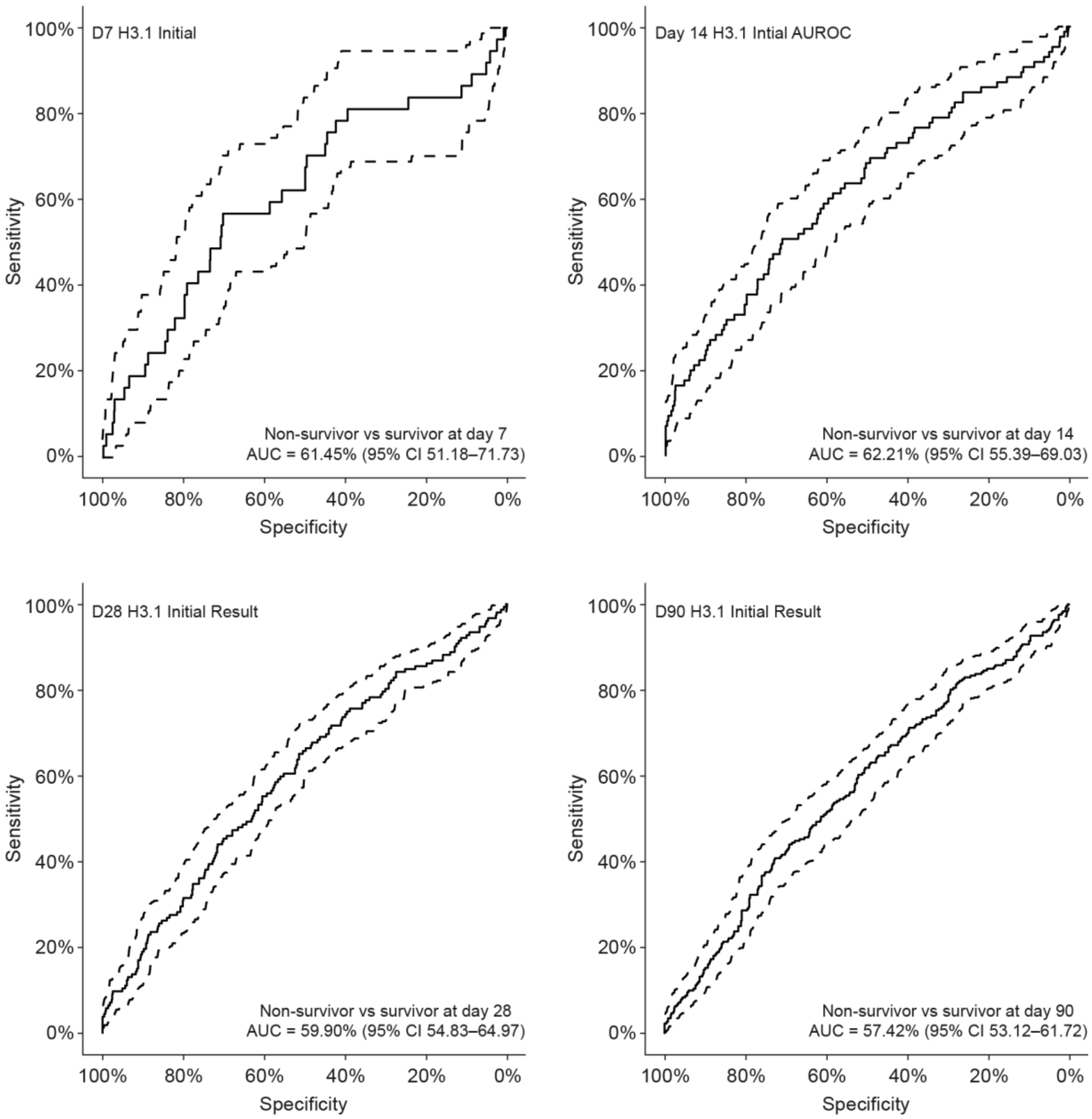

The performance of H3.1 demonstrates moderate discriminatory capability, peaking on day 14 (AUC = 62.21%). Its AUC consistently exceeds 50%, indicating some predictive value, though limited in robustness. H3.1 and lactate exhibit the most consistent moderate predictive capabilities, particularly within the first 2 weeks (days 7 and 14).

WCC shows poor to marginal predictive ability, with its AUC values only slightly above random chance. CRP demonstrates negligible predictive value across all time points. Both H3.1 and lactate, peak discriminatory performance occurs around days 7 and 14, suggesting these markers may have greater utility in the early post-initial phase.

AUC, area under the curve; CI, confidence interval; CRP, C-reactive protein; ROC, receiver operating characteristic; WCC white cell count.

**Fig. 3** Initial WCC day 7, 14, 28 and 90 ROC curves

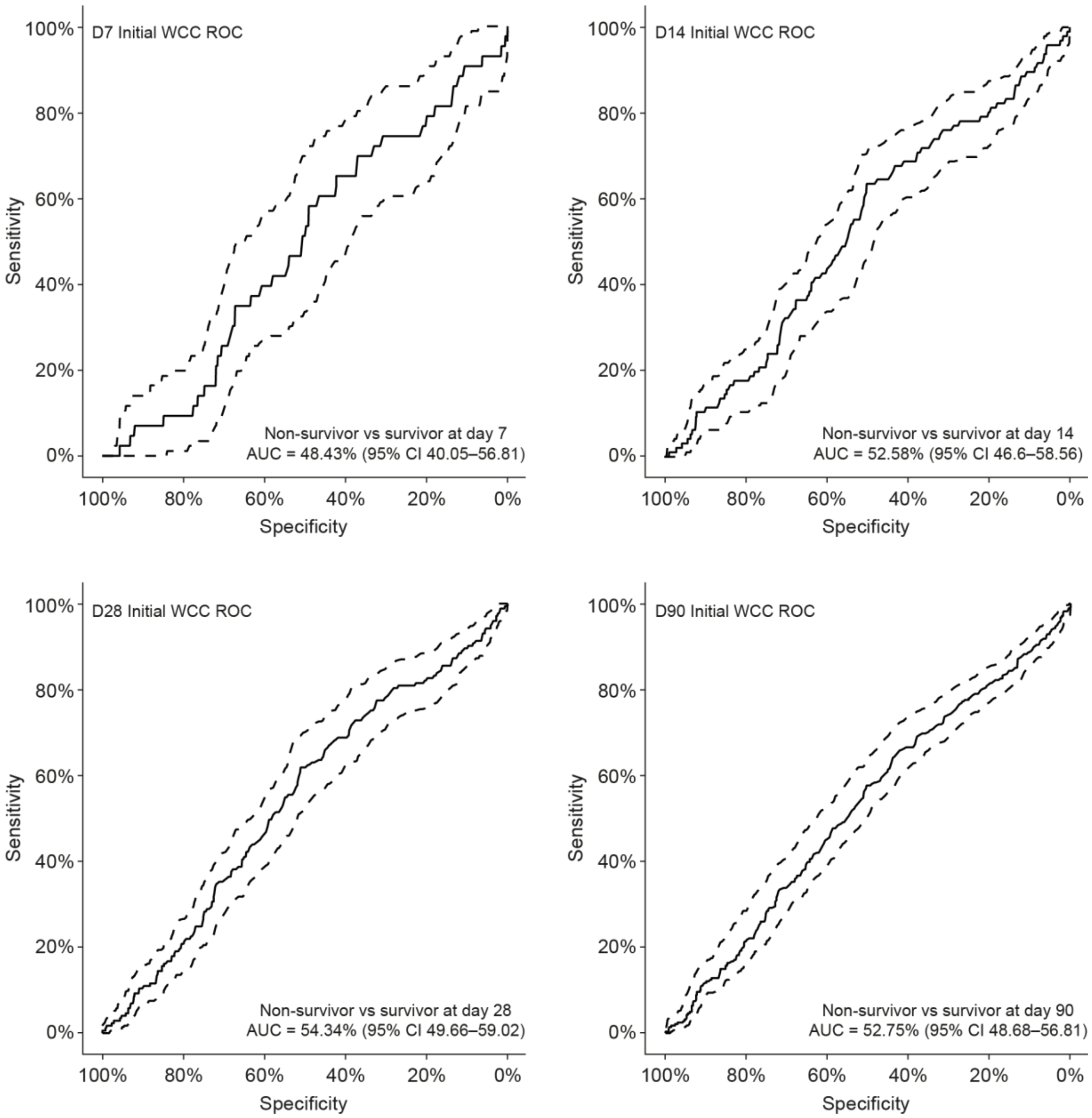

WCC exhibits poor discriminatory power, with AUC values hovering near or below 50%. There is little appreciable change in performance over time.

AUC, area under the curve; CI, confidence interval; ROC, receiver operating characteristic; WCC, white cell count.

**Fig. 4** Maximum lactate day 7, 14, 28 and 90 ROC curves

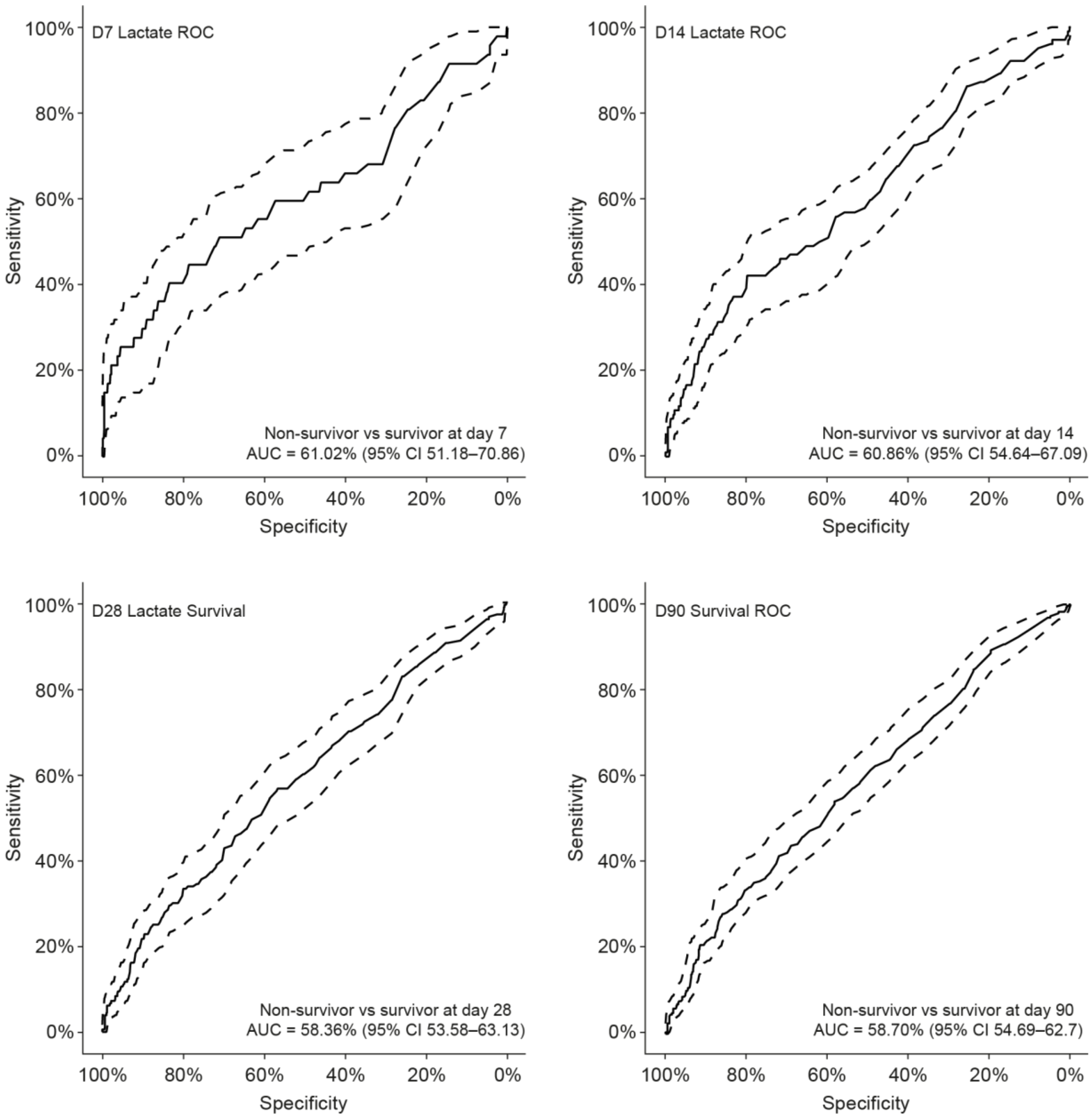

AUC, area under the curve; CI, confidence interval; ROC, receiver operating characteristic.

**Fig. 5** Initial CRP day 7, 14, 28 and 90 ROC curves

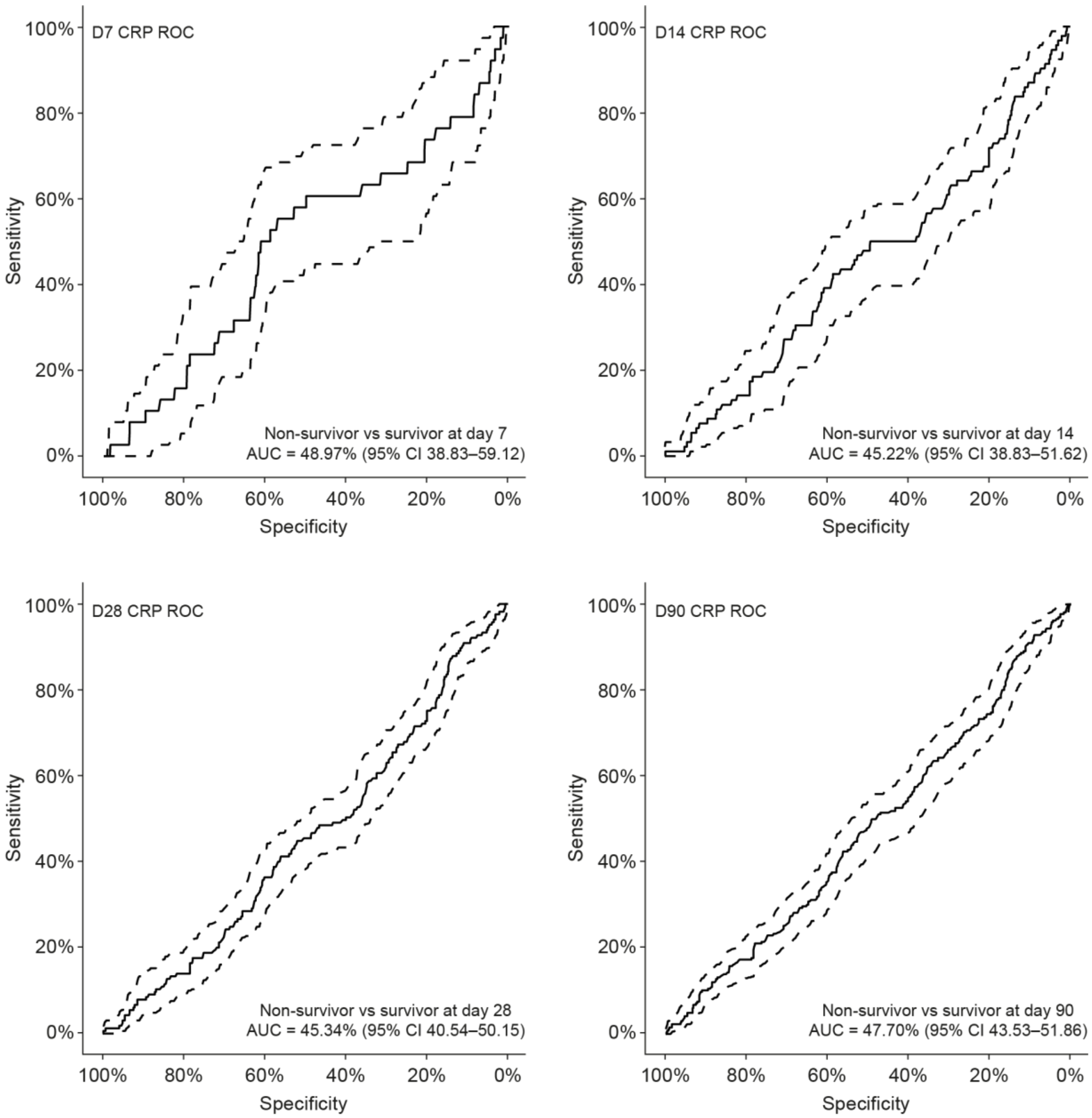

AUC, area under the curve; CI, confidence interval; CRP, C-reactive protein; ROC, receiver operating characteristic.

**Fig. 6** Initial procalcitonin day 7, 14, 28 and 90 ROC curves

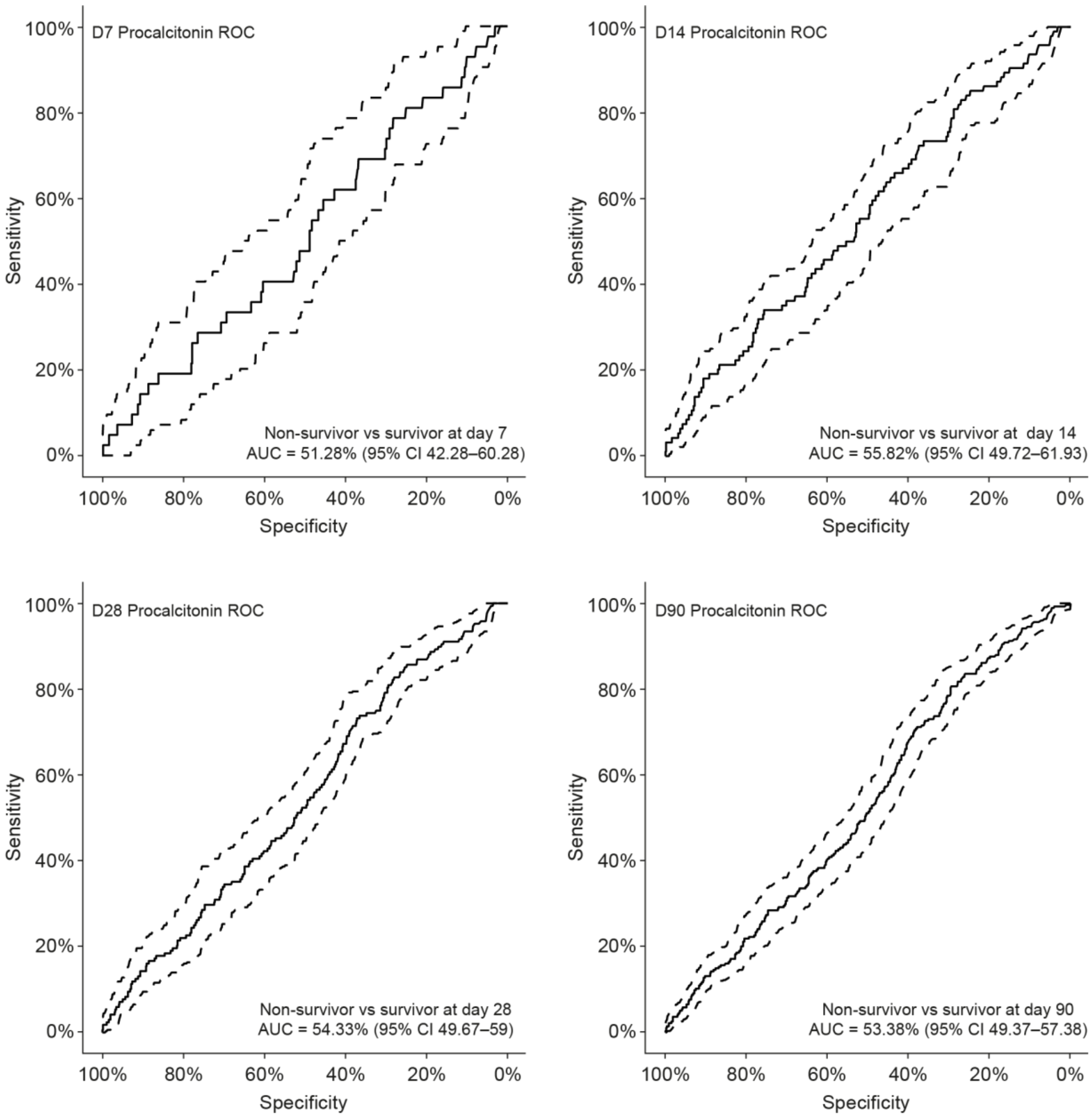

AUC, area under the curve; CI, confidence interval; CRP, C-reactive protein; ROC, receiver operating characteristic.

#### Appendix H: Additional information referenced but content not specified in the manuscript

**Fig. 7** Initial H3.1 readings above 10,000 predict mortality within 14 days

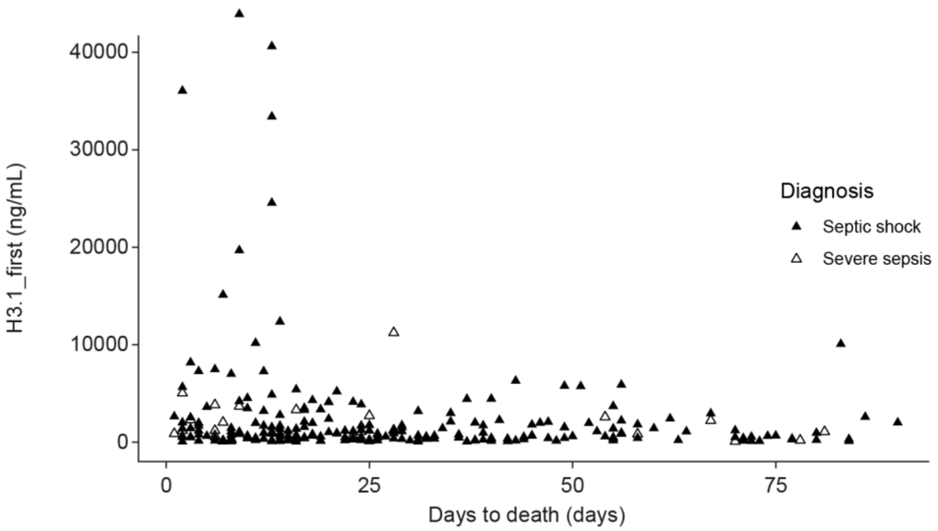

**Table 2** 14-day mortality risk for initial H3.1 nucleosome levels

| H3.1 nucleosomes, ng/mL | Patients, n | | | Mortality risk, % |
|---|---|---|---|---|
| | Total (n=869) | Survivors (n=784) | Died (n=85) | |
| >20,000 | 5 | 0 | 5 | 100 |
| 10,000–20,000 | 16 | 12 | 4 | 25 |
| 1,000–10,000 | 300 | 264 | 36 | 12 |
| <1000 | 548 | 508 | 40 | 7 |

**Fig. 8** Presentation SOFA score and mortality

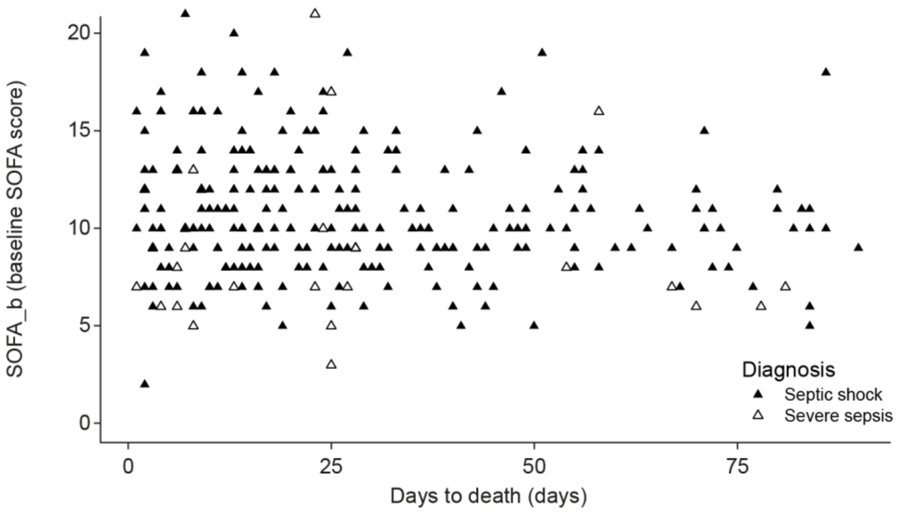

SOFA, Sequential Organ Failure Assessment.

**Table 3** Hazard ratios over time for H3.1

Hazard ratios for H3.1_first_log10

| Measure | Hazard ratio | Lower 95% CI | Upper 95% CI | P value |
|---|---|---|---|---|
| Overall mortality | 1.544 | 1.236 | 1.929 | 0.000129 |
| 7-day mortality | 2.2798 | 1.297 | **4.006** | 0.00417 |
| 14-day mortality | **2.2969** | **1.589** | 3.319 | 9.59e-06 |
| 21-day mortality | 2.1261 | 1.553 | 2.911 | 2.52e-06 |
| 28-day mortality | 1.8641 | 1.408 | 2.468 | 1.37e-05 |
| 35-day mortality | 1.7727 | 1.356 | 2.317 | 2.77e-05 |
| 42-day mortality | 1.6231 | 1.258 | 2.095 | 0.000199 |
| 49-day mortality | 1.6213 | 1.268 | 2.073 | 0.000117 |
| 56-day mortality | 1.6735 | 1.320 | 2.122 | 2.1e-05 |
| 63-day mortality | 1.6721 | 1.323 | 2.113 | 1.68e-05 |
| 70-day mortality | 1.6670 | 1.323 | 2.100 | 1.46e-05 |
| 77-day mortality | 1.5571 | 1.242 | 1.953 | 0.000127 |
| 84-day mortality | 1.5264 | 1.221 | 1.908 | 0.000206 |
| 90-day mortality | 1.5440 | 1.236 | 1.929 | 0.000129 |

CI, confidence interval.

**Table 4** Hazard ratios for 28-day mortality

| Measure | Hazard ratio | Lower 95% CI | Upper 95% CI | P value |
|---|---|---|---|---|
| H3.1_first_log10 | 1.8641 | 1.408 | 2.468 | 1.37e-05 |
| Thrombocytes_first_log10 | 0.3552 | 0.242 | **0.522** | 1.35e-07 |
| Procalcitonin_first_log10 | 1.2307 | 1.014 | 1.493 | 0.0355 |
| Leukocytes_first_log10 | 1.3830 | 0.7717 | 2.478 | 0.276 |
| Lakmax_log10 | **2.6404** | 1.654 | **4.215** | 4.73e-05 |
| Crp_first_log10 | 0.7103 | 0.434 | 1.162 | 0.173 |

CI, confidence interval.

**Table 5** Hazard ratios for all presentation biomarkers and 90-day mortality

| Measure | Hazard ratio | Lower 95% CI | Upper 95% CI | P value |
|---|---|---|---|---|
| H3.1_first_log10 | 1.544 | 1.236 | 1.929 | 0.000129 |
| Thrombocytes_first_log10 | 0.5260 | 0.376 | 0.736 | 0.000177 |
| Procalcitonin_first_log10 | 1.16478 | 1.003 | 1.353 | 0.0458 |
| Leukocytes_first_log10 | 1.1973 | 0.7585 | 1.89 | 0.439 |
| Crp_first_log10 | 0.8787 | 0.584 | 1.322 | 0.535 |

CI, confidence interval.

**Table 6** Optimal survminer cutpoints for different measures of mortality

| **Biomarker**<br>**Units** | **H3.1_first**<br>**ng/mL** | **Crp_first**<br>**mg/L** | **Creatinine_first**<br>**mcmol/L** | **Lakmax_si**<br>**mmol/L** | **Thrombocytes_first**<br>**$10^9$/L** | **Urea_first**<br>**mmol/L** | **Urin24_b**<br>**mL/24h** |
|---|---|---|---|---|---|---|---|
| Overall mortality | 1,142 | 142 | 111 | 6 | 84 | 13 | 1,050 |
| 7-day mortality | 1,149 | 246 | 118 | 11 | 84 | 31 | 15 |
| 14-day mortality | 6,796 | 142 | 159 | 13 | 84 | 16 | 1,036 |
| 21-day mortality | 6,796 | 142 | 114 | 7 | 84 | 16 | 1,036 |
| 28-day mortality | 6,796 | 195 | 111 | 13 | 84 | 16 | 1,036 |
| 35-day mortality | 6,729 | 299 | 111 | 6 | 86 | 16 | 1,036 |
| 42-day mortality | 6,729 | 299 | 111 | 6 | 86 | 13 | 1,036 |
| 49-day mortality | 6,427 | 299 | 114 | 6 | 86 | 16 | 1,036 |
| 56-day mortality | 1,142 | 299 | 111 | 6 | 86 | 13 | 1,036 |
| 63-day mortality | 1,142 | 299 | 111 | 6 | 86 | 13 | 1,036 |
| 70-day mortality | 1,142 | 299 | 111 | 6 | 86 | 12 | 1,043 |
| 77-day mortality | 1,142 | 299 | 111 | 6 | 86 | 13 | 1,043 |
| 84-day mortality | 1,142 | 142 | 111 | 6 | 86 | 13 | 1,043 |
| 90-day mortality | 1,142 | 142 | 111 | 6 | 84 | 13 | 1,050 |

**Fig. 9** CONSORT diagram explaining the analysis of AKI

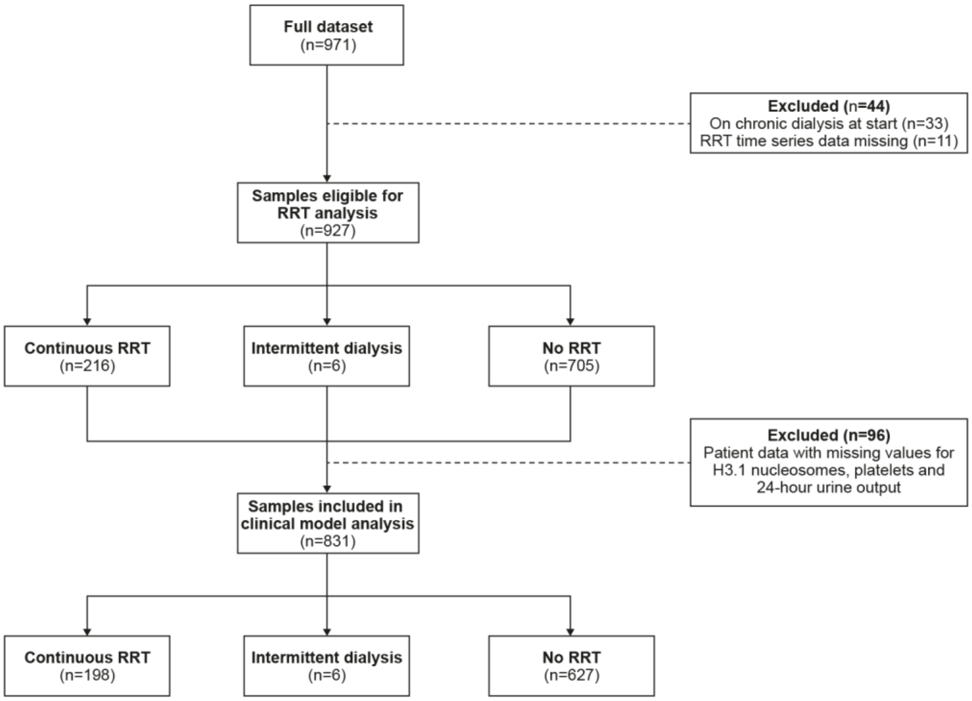

AKI, acute kidney injury; RRT, renal replacement therapy.

**Fig. 10** Correlation between initial H3.1 and initial creatinine

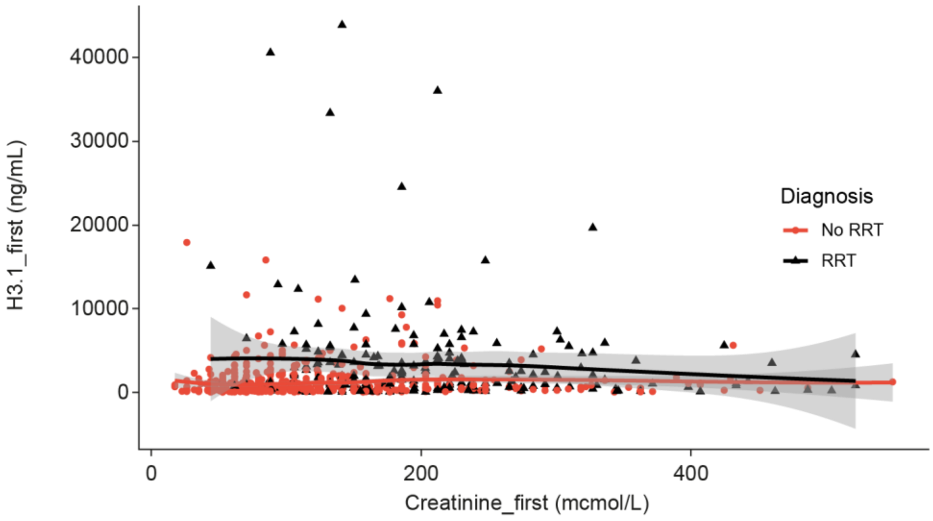

This scatterplot illustrates the relationship between initial serum creatinine and initial H3.1 levels in patients stratified by renal replacement therapy (RRT) status. The analysis reveals modest correlations between these parameters, with a Pearson correlation coefficient of 9% and a Spearman correlation coefficient of 23%, suggesting a weak positive relationship. The regression lines indicate a slight negative trend for both groups across increasing creatinine levels, though the wide confidence intervals at higher creatinine values suggest considerable uncertainty in this relationship. The distribution pattern suggests that while there may be some association between renal dysfunction (as indicated by creatinine levels) and H3.1 concentrations, other factors likely contribute to the substantial variability observed in H3.1 levels, particularly among patients requiring RRT.

**Fig. 11** Predictive performance of creatinine, H3.1 nucleosomes, CRP, lactate, WCC, platelet count and urine output for renal replacement therapy

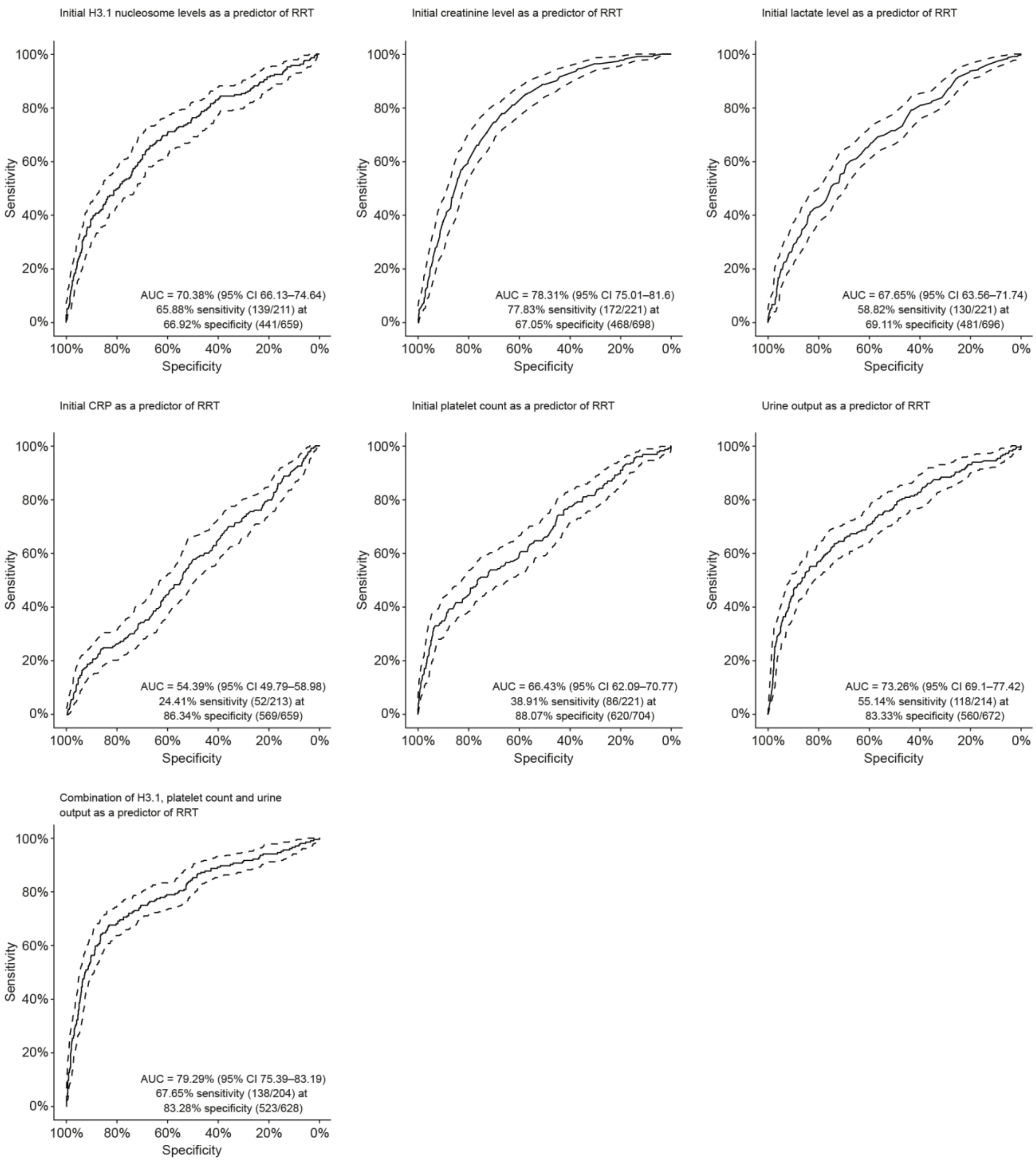

AUC, area under the curve; CI, confidence interval; CRP, C-reactive protein; ROC, receiver operating characteristic; RRT, renal replacement therapy; WCC, white cell count.

**Fig. 12** Clinical model of H3.1 + platelets + urine 24-h for anyRRT28

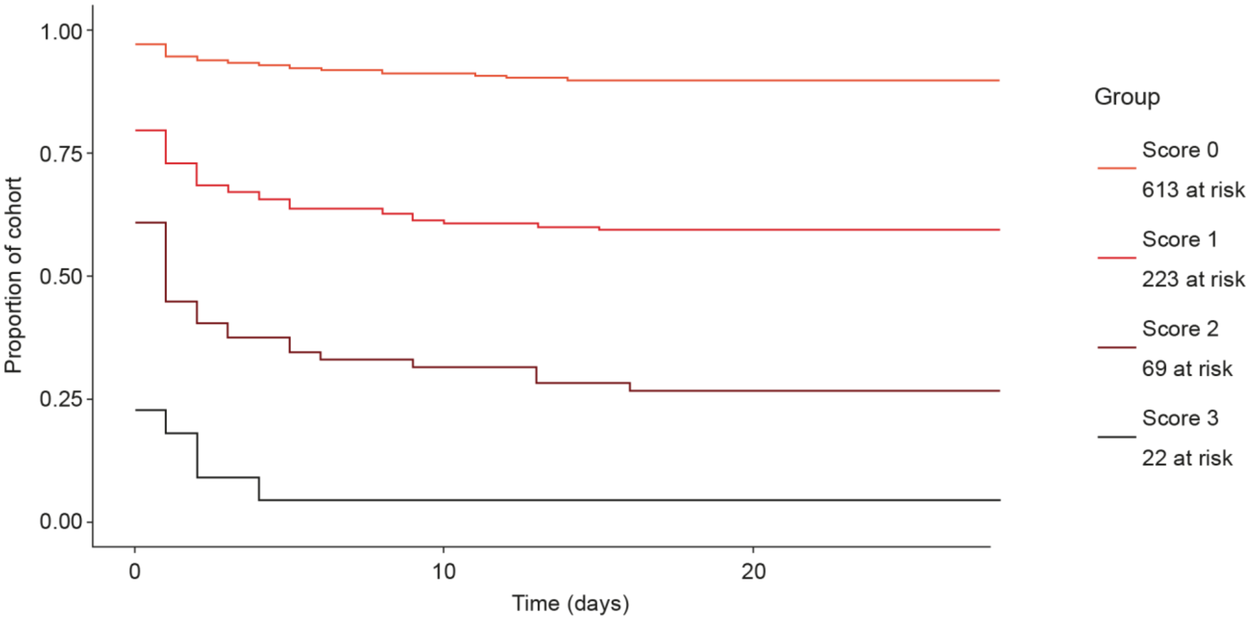

A Kaplan–Meier analysis showing RRT-free survival over 28 days stratified by clinical risk scores. Scores were calculated based on admission H3.1 nucleosome levels >2500 ng/mL, platelet count <84x$10^9$/L, and 24-hour urine output <28 mL/kg/hr. Each parameter contributed one point to the total score (range 0–3). The cohort (n=927) was divided into four groups: Score 0 (n=613), Score 1 (n=223), Score 2 (n=69), and Score 3 (n=22). Higher scores were associated with progressively worse RRT-free survival.

RRT, renal replacement therapy.

**Fig. 13** Linear regression analysis between biomarker levels and RRT a) H3.1 nucleosome, b) platelets, c) WBC, d) CRP and e) procalcitonin

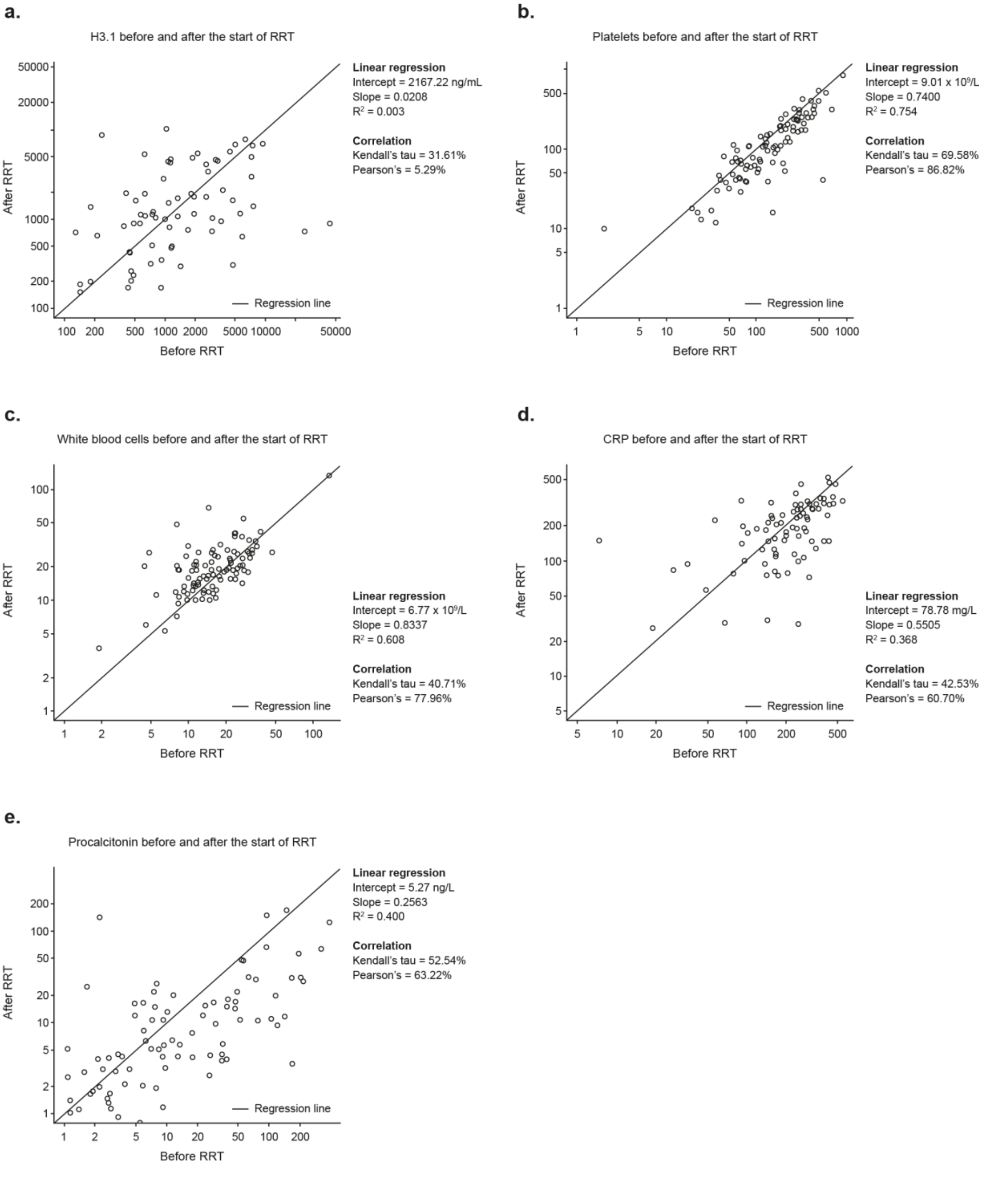

This figure presents linear regression analyses examining the relationships between various biomarkers before and after the start of RRT: a) H3.1 nucleosome shows weak correlation ($R^2$=0.023, Pearson's=0.29%) with moderate slope (0.0006); b) platelets demonstrate strong correlation ($R^2$=0.754, Pearson's=86.02%) with positive slope (0.7400); c) WBCs show moderate correlation ($R^2$=0.038, Pearson's=77.06%) with slope of 0.8037; d) CRP exhibits moderate correlation ($R^2$=0.366, Pearson's=60.70%) with slope of 0.5505; and e) procalcitonin displays moderate correlation ($R^2$=0.400, Pearson's=63.29%) with slope of 0.2563.

CRP, C-reactive protein; R, coefficient of determination; RRT, renal replacement therapy; WBC, white blood cells.

**Fig. 14** Hazard ratios for log-transformed initial H3.1 measurement for renal replacement therapy (RRT)

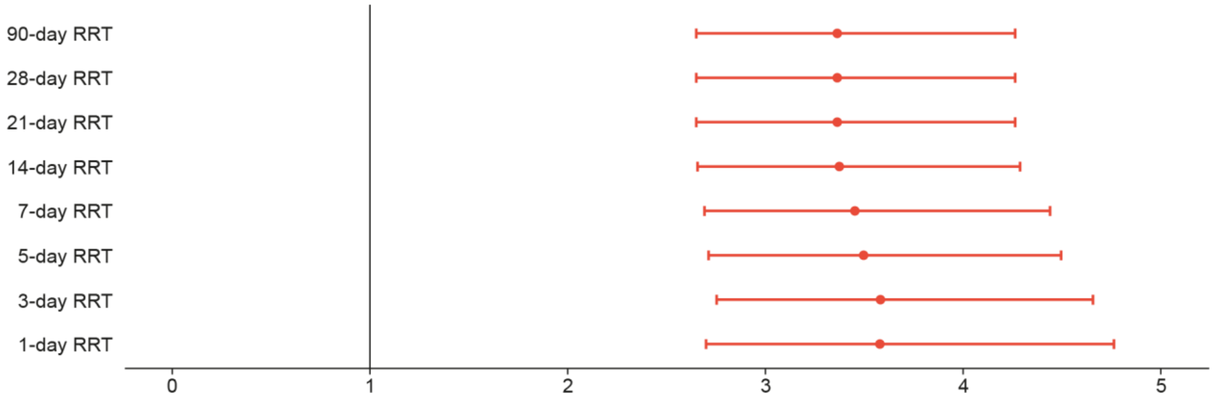

**Table 7** Hazard ratios for log-transformed initial H3.1 measurement for renal replacement therapy (RRT)

| H3.1_first_log10 | HR estimate | CI: lower 95% | CI: Upper 95% |
|---|---|---|---|
| 1-day RRT | 3.575 | 2.689 | 4.753 |
| 3-day RRT | 3.5788 | 2.751 | 4.655 |
| 5-day RRT | 3.4903 | 2.709 | 4.496 |
| 7-day RRT | 3.4528 | 2.686 | 4.438 |
| 14-day RRT | 3.3715 | 2.652 | 4.287 |
| 21-day RRT | 3.3591 | 2.646 | 4.264 |
| 28-day RRT | 3.3591 | 2.646 | 4.264 |
| 90-day RRT | 3.3591 | 2.646 | 4.264 |

CI, confidence interval; HR, hazard ratio; RRT, renal replacement therapy.
